# Cardiometabolic risk factors associated with brain age and accelerate brain ageing

**DOI:** 10.1101/2021.02.25.21252272

**Authors:** Dani Beck, Ann-Marie G. de Lange, Mads L. Pedersen, Dag Alnæs, Ivan I. Maximov, Irene Voldsbekk, Geneviève Richard, Anne-Marthe Sanders, Kristine M. Ulrichsen, Erlend S. Dørum, Knut K. Kolskår, Einar A. Høgestøl, Nils Eiel Steen, Srdjan Djurovic, Ole A. Andreassen, Jan E. Nordvik, Tobias Kaufmann, Lars T. Westlye

## Abstract

The structure and integrity of the ageing brain is interchangeably linked to physical health, and cardiometabolic risk factors (CMRs) are associated with dementia and other brain disorders. In this mixed cross-sectional and longitudinal study (interval mean and standard deviation = 19.7 ± 0.5 months), including 1062 datasets from 790 healthy individuals (mean (range) age = 46.7 (18-94) years, 54% women), we investigated CMRs and health indicators including anthropometric measures, lifestyle factors, and blood biomarkers in relation to brain structure using MRI-based morphometry and diffusion tensor imaging (DTI). We performed tissue specific brain age prediction using machine learning and performed Bayesian multilevel modelling to assess changes in each CMR over time, their respective association with brain age gap (BAG), and their interaction effects with time and age on the tissue-specific BAGs. The results showed credible associations between DTI-based BAG and blood levels of phosphate and mean cell volume (MCV), and between T1-based BAG and systolic blood pressure, smoking, pulse, and C-reactive protein (CRP), indicating older-appearing brains in people with higher cardiometabolic risk (smoking, higher blood pressure and pulse, low-grade inflammation). Longitudinal evidence supported interactions between both BAGs and waist-to-hip ratio (WHR), and between DTI-based BAG and systolic blood pressure and smoking, indicating accelerated ageing in people with higher cardiometabolic risk (smoking, higher blood pressure, and WHR). The results demonstrate that cardiometabolic risk factors are associated with brain ageing. While randomised controlled trials are needed to establish causality, our results indicate that public health initiatives and treatment strategies targeting modifiable cardiometabolic risk factors may also improve risk trajectories and delay brain ageing.

## 1. Introduction

It is well established that various CMRs are associated with increased risk of a range of brain disorders, including stroke, Alzheimer’s disease and other dementias, in addition to ageing-related cognitive decline, supporting an intimate body–brain connection in ageing (Qiu & Fratiglioni, 2015). Moreover, associations between high insulin and obesity in childhood and risk for psychosis and depression at 24 years of age indicate that CMRs in childhood represent predictors for mental disorders later in life (Perry et al., 2021). Research has found that established CMRs such as blood pressure (Fuhrmann et al., 2019; Verhaaren et al., 2013), WHR, body mass index (BMI) (Karlsson et al., 2013; Spangaro et al., 2018), diabetes mellitus (Hoogenboom et al., 2014; Hsu et al., 2012), hypertension (McEvoy et al., 2015), total elevated cholesterol (Walhovd et al., 2014; Williams et al., 2018), smoking (Jeerakathil et al., 2004), and high low-density lipoprotein (LDL) cholesterol (Murray et al., 2005), are all associated with brain structure to various degrees. However, there is substantial variability among individuals in terms of impact on the brain and the putative biological factors involved.

Brain-predicted age has recently emerged as a reliable and heritable biomarker of brain health and ageing (Cole et al., 2017; Franke et al., 2010; Kaufmann et al., 2019). The difference between the brain-predicted age and chronological age - also referred to as the brain age gap (BAG) - can be used to assess deviations from expected age trajectories. These estimations of brain age may thus have clinical implications, as identifying factors associated with higher BAG and accelerated ageing can help us detect potential targets for intervention strategies.

Higher brain age has been associated with poorer cognitive functioning in healthy individuals (Richard et al., 2018) and people with cognitive impairment (Varatharajah et al., 2018), mild cognitive impairment (MCI), dementia (Kaufmann et al., 2019), and mortality in elderly people (Cole et al., 2018). Larger BAGs have also been reported among patients with psychiatric and neurological disorders, including schizophrenia, bipolar disorder, multiple sclerosis (Høgestøl et al., 2019; Kaufmann et al., 2019; Tønnesen et al., 2020), depression (Han et al., 2020), and epilepsy (Pardoe et al., 2017; Sone et al., 2019).

While BAG shows substantial heritability (Cole et al., 2017; Kaufmann, et al., 2019), the rate of brain ageing is malleable and dependent on a range of life events and health and lifestyle factors (Cole, 2020; Lindenberger, 2014; Sanders et al., 2021). Understanding the impact of cardiometabolic risk on brain integrity and ageing represents a window of opportunity wherein interventions targeting key elements of cardiometabolic health may delay and even prevent pathological brain changes (Friedman et al., 2014).

Studies assessing cardiometabolic risk have reported brain age associations with diastolic blood pressure, BMI (Franke et al., 2014), obesity (Kolenic et al., 2018; Ronan et al., 2016), and diabetes (Franke et al. 2013). Larger BAGs have also been associated high blood pressure, alcohol intake, diabetes, smoking, and history of stroke in the UK Biobank (Cole, 2020), and with high blood pressure, alcohol intake, and stroke risk scores in the Whitehall II MRI sub-sample (de Lange et al. 2020). Despite existing research, the links between cardiometabolic risk and brain ageing are still unclear. Longitudinal studies utilising multimodal imaging may aid to link individual CMRs to tissue specific effects.

By including cross-sectional and longitudinal data obtained from 790 healthy subjects aged 18-94 years (mean 46.7, SD 16.3), our primary aim was to investigate how key CMRs interact with tissue-specific (DTI and T1-weighted) measures of brain ageing. We investigated longitudinal associations between brain age and a range of CMRs and tested both for main effects across time and interactions with age and time. Adopting a Bayesian statistical framework, we hypothesized that key indicators of cardiometabolic risk would be associated with more apparent brain ageing, both reflected as main effects across time, and as interactions, indicating a faster pace of brain ageing over the course of the follow-up period in people with high cardiometabolic risk.

## 2. Material and methods

### 2.1. Sample description

The initial sample consisted of 1130 (832 baseline, 298 follow up) datasets from 832 healthy participants from two integrated studies; the Thematically Organised Psychosis (TOP) (Tønnesen et al., 2018) and StrokeMRI (Richard et al., 2018). Exclusion criteria included neurological and mental disorders, and previous head trauma. The study was conducted in line with the Declaration of Helsinki and approved by the Regional Ethics Committee, and all participants provided written informed consent. Following the removal of 68 MRI datasets after quality checking (QC) of the MRI data (see section 2.5), the final sample comprised 1062 datasets from 790 individuals, including longitudinal data (two time-points with 19.7 months interval on average (min = 9.8, max = 35.6) from 272 participants. Demographic information of the test sample is summarised in Table 1, Figure 1.

**Figure 1.**
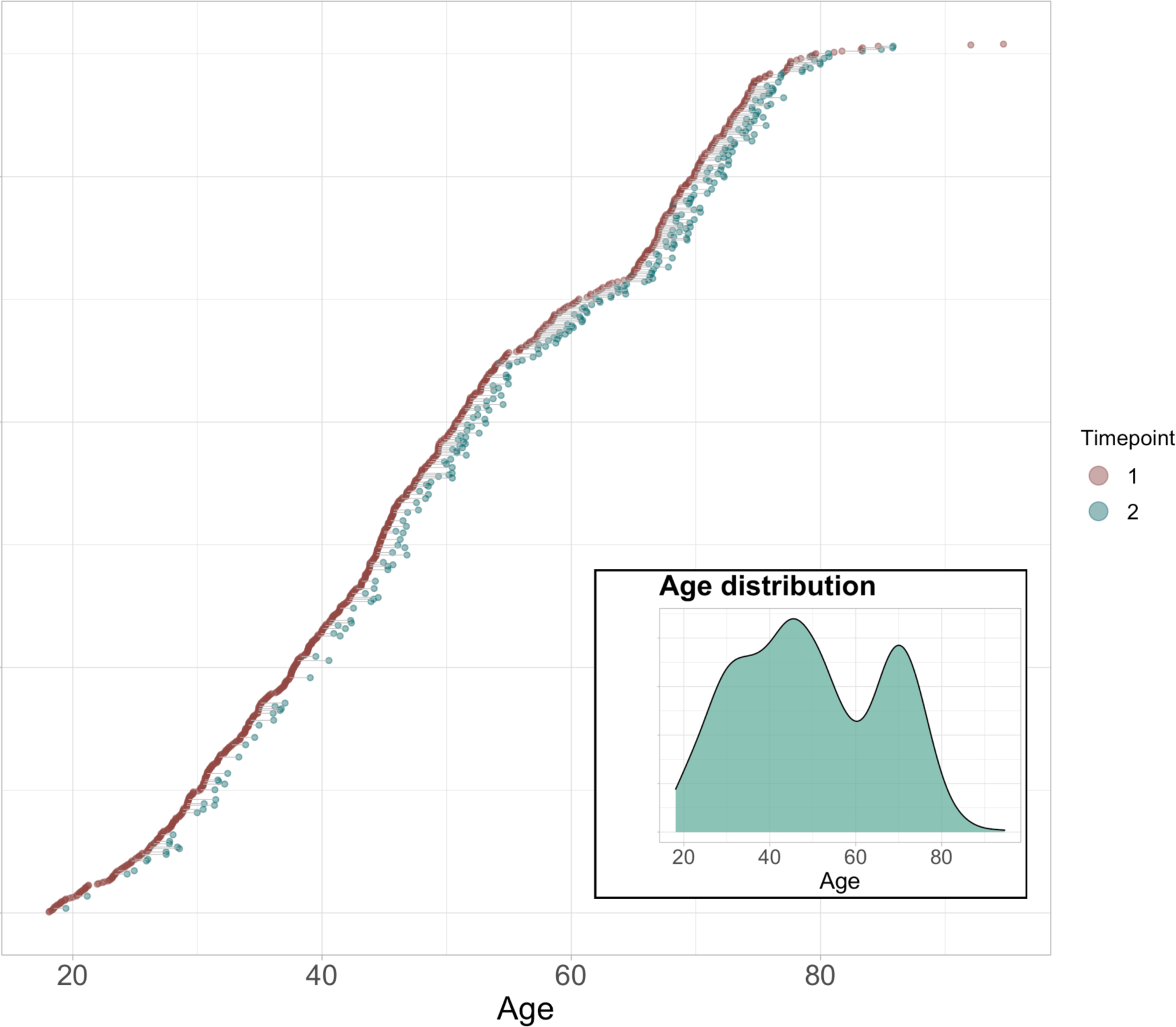
Available baseline and follow-up data. All participants are shown. Participants with data at baseline are visualised in red dots (N = 790). The subset (n = 272) with longitudinal measures are connected to corresponding timepoint with green dots. The mean interval between timepoints was 1.64 years (SD = 0.5 years). Subplot shows age distribution at baseline.

**Table 1.**
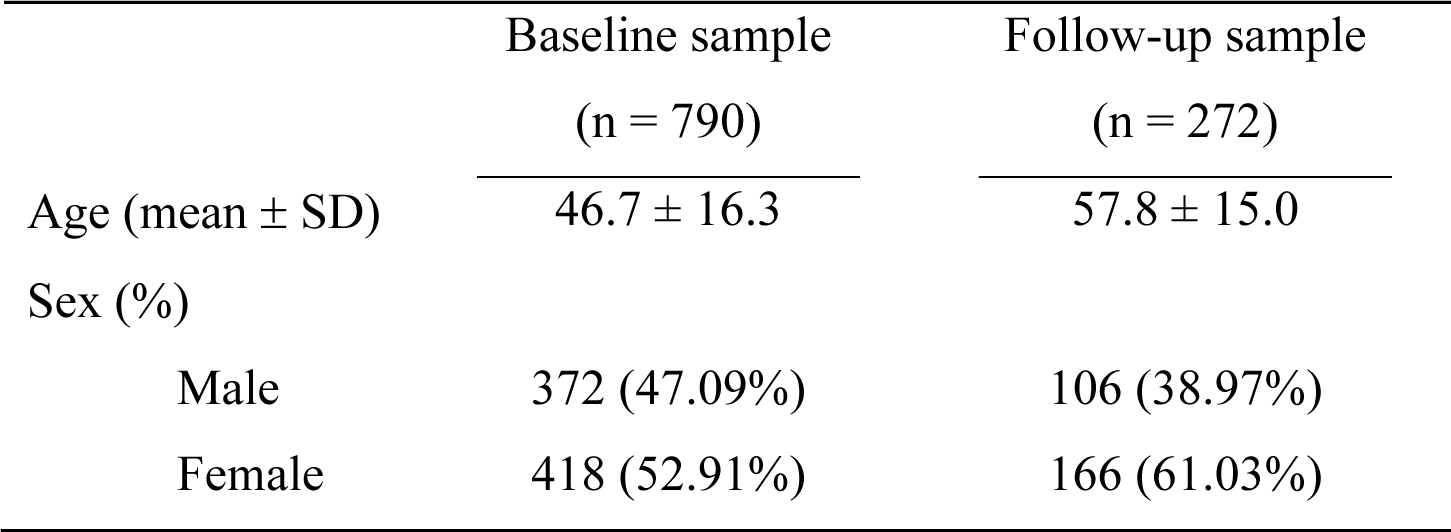
Sample descriptives at baseline and follow-up.

|  | Baseline sample<br>(n = 790) | Follow-up sample<br>(n = 272) |
| --- | --- | --- |
| Age (mean ± SD) | $46.7 \pm 16.3$ | $57.8 \pm 15.0$ |
| Sex (%) |  |  |
| Male | 372 (47.09%) | 106 (38.97%) |
| Female | 418 (52.91%) | 166 (61.03%) |

Data from the Cambridge Centre for Ageing and Neuroscience (Cam-CAN: http://www.mrc-cbu.cam.ac.uk/datasets/camcan/; Shafto et al., 2014; Taylor et al., 2017) was used as an independent training sample for brain age prediction (see section 2.6). After QC, MRI data from 622 participants were included (age range = 18–87, mean age ± standard deviation = 54.2 ± 18.4). SI Figure 1 shows the age distribution for the training and test samples.

### 2.2. MRI acquisition

MRI was performed at Oslo University Hospital on a GE Discovery MR750 3T scanner with a 32-channel head coil. DTI data were acquired with a spin echo planar imaging (EPI) sequence with the following parameters: repetition time (TR)/echo time (TE)/flip angle: 8,150 ms/83.1 ms/90^◦^, FOV: 256 × 256 mm^2^, slice thickness: 2 mm, in-plane resolution: 2×2 mm^2^, 60 non-coplanar directions (b = 1000 s/mm^2^) and 5 b = 0 volumes, scan time: 8:58 min. In addition, 7 b = 0 volumes with reversed phase-encoding direction were acquired. High-resolution T1-weighted data was acquired using a 3D inversion recovery prepared fast spoiled gradient recalled sequence (IR-FSPGR; BRAVO) with the following parameters: TR: 8.16 ms, TE: 3.18 ms, flip angle: 12^◦^, voxel size: 1×1×1 mm^3^, FOV: 256 × 256 mm^2^, 188 sagittal slices, scan time: 4:43 min.

For the Cam-CAN training set, participants were scanned on a 3T Siemens TIM Trio scanner with a 32-channel head-coil at Medical Research Council (UK) Cognition and Brain Sciences Unit (MRC-CBSU) in Cambridge, UK. DTI data was acquired using a twice— refocused spin echo sequence with the following parameters a TR: 9,100 ms, TE: 104 ms, FOV: 192 × 192 mm^2^, voxel size: 2 mm, 66 axial slices using 30 directions with b = 1000 s/mm^2^, 30 directions with b = 2000 s/mm^2^, and 3 b = 0 images (Shafto et al., 2014). High-resolution 3D T1-weighted data was acquired using a magnetisation prepared rapid gradient echo (MPRAGE) sequence with the following parameters: TR: 2,250 ms, TE: 2.99 ms, inversion time (TI): 900 ms, flip angle: 9^◦^, FOV of 256 × 240 × 192 mm^3^; voxel size = 1×1×1 mm^3^, GRAPPA acceleration factor of 2, scan time 4:32 min (Shafto et al., 2014).

### 2.3. DTI processing and TBSS analysis

Processing steps for single-shell DTI data in the test set followed a previously described pipeline (Maximov et al., 2019), including noise correction (Veraart et al., 2016), Gibbs ringing correction (Kellner et al., 2016), corrections for susceptibility induced distortions, head movements and eddy current induced distortions using topup (http://fsl.fmrib.ox.ac.uk/fsl/fslwiki/topup) and eddy (http://fsl.fmrib.ox.ac.uk/fsl/fslwiki/eddy) (Andersson & Sotiropoulos, 2016). Isotropic smoothing was carried out with a Gaussian kernel of 1 mm^3^ implemented in the FSL function *fslmaths.* DTI metrics were estimated using *dtifit* in FSL and a weighted least squares algorithm. Processing steps for the training set followed a similar pipeline with the exception of the noise correction procedure. Voxelwise statistical analysis of the fractional anisotropy (FA) data was carried out using Tract-Based Spatial Statistics (TBSS) (Smith et al., 2006), as part of FSL (Smith et al., 2004). First, FA images were brain-extracted using BET (Smith, 2002) and aligned into a common space (FMRI58_FA template) using the nonlinear registration tool FNIRT (Andersson, Jenkinson, & Smith., 2007; Jenkinson et al., 2012), which uses a b-spline representation of the registration warp field (Rueckert et al., 1999). Next, the mean FA image of all subjects was created and thinned to create a mean FA skeleton that represents the centres of all tracts common to the group. Each subject’s aligned FA data was then projected onto this skeleton.

The mean FA skeleton was thresholded at FA > 0.2. This procedure was repeated in order to extract axial diffusivity (AD), mean diffusivity (MD), and radial diffusivity (RD). *fslmeants* was used to extract the mean skeleton and 20 regions of interest (ROI) based on a probabilistic white matter atlas (JHU) (Hua et al., 2008) for each metric. Including the mean skeleton values, 276 features per individual were derived in total.

### 2.4. FreeSurfer processing

T1-weighted MRI data were processed using FreeSurfer (Fischl, 2012) 7.1.0 for the test set and FreeSurfer 5.3 for the training set. To extract reliable area, volume and thickness estimates, the test set including follow-up data were processed with the longitudinal stream (Reuter et al., 2012) in FreeSurfer. Specifically, an unbiased within-subject template space and image (Reuter & Fischl, 2011) is created using robust, inverse consistent registration (Reuter et al., 2010). Several processing steps, such as skull stripping, Talairach transforms, atlas registration as well as spherical surface maps and parcellations are then initialized with common information from the within-subject template, significantly increasing reliability and statistical power (Reuter et al., 2012). Due to the longitudinal stream in FreeSurfer influencing the thickness estimates, and subsequently having an impact on brain age prediction (Høgestøl et al., 2019), both cross-sectional and longitudinal data in the test set were processed with the longitudinal stream. Cortical parcellation was performed using the Desikan-Killiany atlas (Desikan et al., 2006), and subcortical segmentation was performed using a probabilistic atlas (Fischl et al., 2002). 269 FreeSurfer based features were extracted in total, including global features for intracranial volume, total surface area, and whole cortex mean thickness, as well as the volume of subcortical structures.

### 2.5. Quality checking procedure

Prior to statistical analyses, a rigorous QC procedure was implemented to ensure sufficient data quality.

For DTI data (N = 1130) we derived various QC metrics (see SI table 1), including temporal signal-to-noise-ratio (tSNR) (Roalf et al., 2016). Datasets with tSNR z > 2.5 standard deviations from the mean were flagged and manually checked and removed if deemed to have unsatisfactory data quality. A total of 14 datasets were removed during QC, leaving the dataset at n = 1116 scans.

For T1-weighted data, QC was carried out using the ENIGMA cortical QC protocol including three major steps: outlier detection, internal surface method, and external surface method. Quality ratings of each image were recorded using the ENIGMA cortical QC template for each of the initial 1130 dataset. A total of 16 datasets were removed, leaving the dataset at n = 1114 scans. Next, the separate datasets from both T1 (N = 1114) and DTI (N = 1116) were merged to form a matching sample by subject ID, leaving the sample at N = 1101, consisting of the same subjects that had quality checked data for both modalities. Finally, this sample was merged with the CMR data, leaving the final sample used for the study at N = 1062.

### 2.6 Brain age prediction

In line with previous studies (Kuhn et al., 2018; Richard et al., 2018), we used Cam-CAN to train the brain age prediction models. The model input included 276 features for the DTI-based age prediction and 269 features for the age prediction based on T1-weighted data, as described in sections 2.3 and 2.4. Age prediction was performed using XGBoost regression (https://xgboost.readthedocs.io/en/latest/python), which is based on a decision-tree ensemble algorithm used in several recent brain age prediction studies (Beck et al., 2021; de Lange et al., 2019; de Lange et al., 2020; Kaufmann et al., 2019; Richard et al., 2020). Parameters were tuned in nested cross-validations using 5 inner folds for grid search, and 10 outer folds for validating model performance within the training sample. Next, the optimised models were applied to the test sample, and R^2^, RMSE, and MAE were calculated to evaluate prediction accuracy in the test set. To adjust for a commonly observed age-bias (overestimated predictions for younger participants and underestimated predictions for older participants) (Liang et al., 2019), we applied a statistical correction by first fitting Y = α × Ω + β, where Y is the modelled predicted age as a function of chronological age (Ω), and *α* and *β* represent the slope and intercept. Next, we used the derived values of *α* and *β* to correct predicted age with *Corrected Predicted Age* = *Predicted Age* + [Ω − (α × Ω + β)] (de Lange & Cole, 2020) before re-calculating R^2^, RMSE, and MAE. BAG was calculated using (corrected predicted age - chronological age) for each of the models, providing T1 and DTI-based BAG values for all participants.

### 2.7. Cardiometabolic risk factors

Clinical information including BMI, systolic and diastolic blood pressure, pulse, WHR, and smoking were collected at the time of MRI, with standard hospital biochemical blood measures being collected at a different site (SI Table 2). All participants underwent a physical examination. BMI (weight in kg/height in m^2^) was calculated from weighing the participants on calibrated digital weights wearing light clothing and no shoes. Waist circumference was measured midway between lowest rib and the iliac crest. Blood pressure was recorded in sitting position after resting before MRI scans were collected and after. Blood samples were drawn and analysed for haemoglobin, erythrocyte indexes (MCV [mean corpuscular volume], MCH [mean corpuscular haemoglobin], MCHC [mean corpuscular haemoglobin concentration]), thrombocytes, sodium, potassium, chloride, calcium, magnesium, phosphate, creatinine, ALAT (alanine transaminase), CK (creatine kinase), LD (lactate dehydrogenase), GT (gamma-glutamyl transferase), CRP (C-reactive protein), total cholesterol, LDL (low-density lipoprotein) cholesterol, HDL (high-density lipoprotein) cholesterol, triglycerides, and glucose. Blood samples were analysed at the Department of Medical Biochemistry, Oslo University Hospital, on several routine instruments: Integra 800, Abbot Architect, i2000, Cobas 8000 e602 and Cobas 8000 e801 (Roche Diagnostics, Basel, Switzerland: www.roche.com/about/business/diagnostics.html) using standard methods controlled by internal and external quality control samples (Rødevand et al., 2019).

**Table 2.** Average R^2^, root mean square error (RMSE), and mean absolute error (MAE) ± standard deviation for the age prediction models within the training sample (Cam-CAN), test set, and age-corrected test set.

|  |  | Training sample | Test set before | Test set after |
| --- | --- | --- | --- | --- |
|  |  | (Cam-CAN) | age-bias correction | age-bias correction |
| DTI | R<sup>2</sup> | 0.82 ± 0.04 | 0.72 | 0.92 |
|  | RMSE | 7.67 ± 0.83 | 10.11 | 5.12 |
|  | MAE | 6.15 ± 0.55 | 8.37 | 4.06 |
| T1 | R<sup>2</sup> | 0.81 ± 0.04 | 0.73 | 0.87 |
|  | RMSE | 7.93 ± 0.84 | 9.11 | 6.55 |
|  | MAE | 6.19 ± 0.83 | 7.2 | 5.21 |

Missing entries (<15% for each variable) were imputed using the MICE package (van Buuren & Groothuis-Oudshoorn, 2011) in R, where five imputations were carried out using the predictive mean matching method (package default). The distribution of the original and imputed data was inspected (SI Figures 2-5) and the imputed data were deemed as plausible values. Of the five imputations, the first was used for the remainder of the study. Additional QC was carried out on all CMRs using a multivariate outlier detection algorithm, where anomalies in the data are detected as observations that do not conform to an expected pattern to other items. Using the R package *mvoutlier* (Filzmoser et al., 2005), potential outliers were flagged using the Mahalanobis distance (SI Figures 6 and 7). Informed by an interactive plot using the *chisq.plot* function, manual outlier observations of each of these flagged values deemed 8 of them as true outliers (SI Figure 8), leading to their removal from the initial 1120 CMR dataset, and leaving the dataset at 1112. The final sample was further reduced to 1062 datasets (from 790 individuals) when merged with the available MRI datasets (N = 1101).

**Figure 2.**
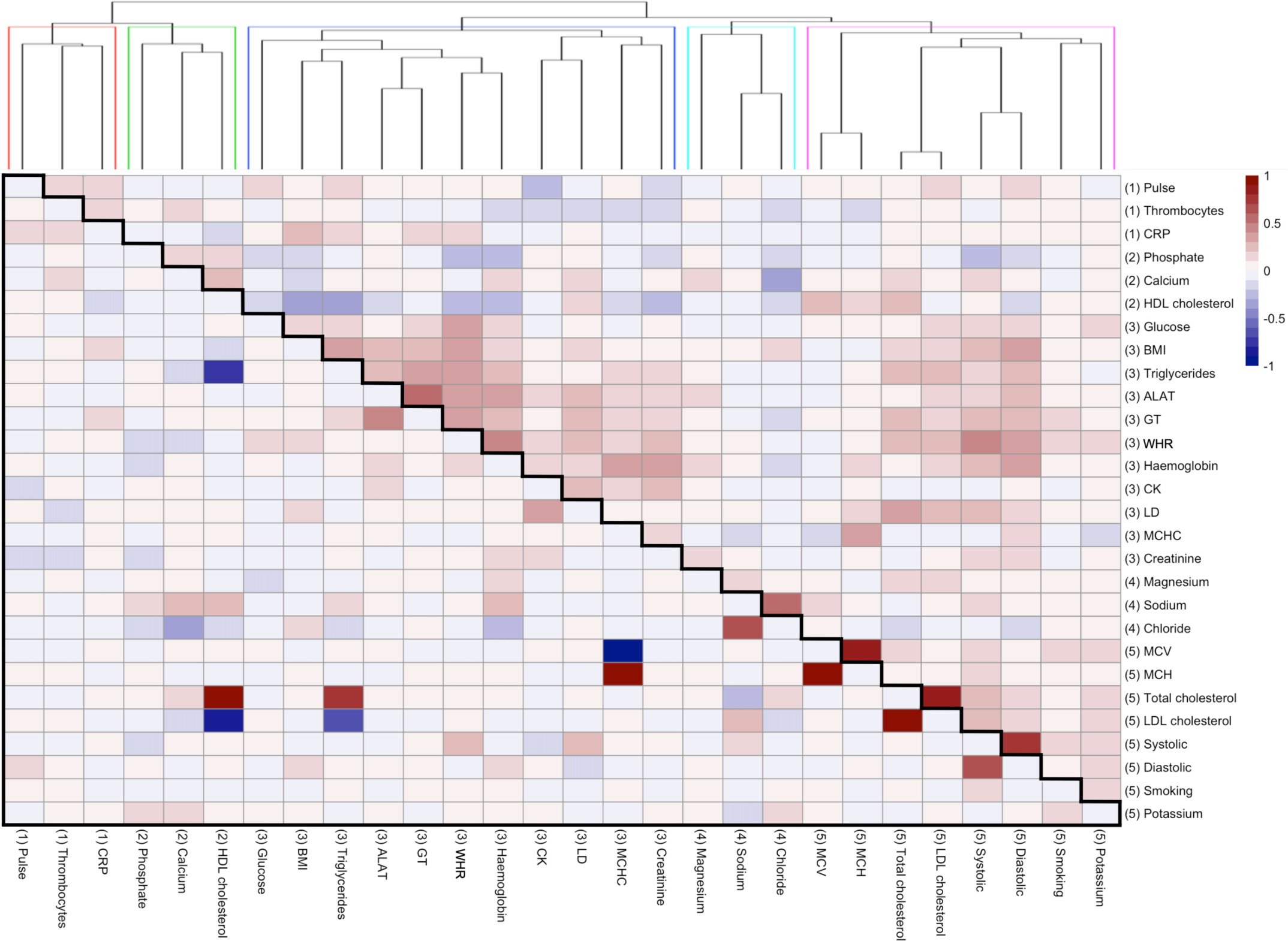
Associations between CMRs. Heatmap showing correlation matrix of all CMRs (scaled), where the lower diagonal shows partial correlations, and the upper diagonal shows full correlations. Hierarchical clustering of the variables was performed based on the full correlations and revealed five cluster groups, shown by numbers in brackets and the higher panel dendrogram split by colour. SI Table 2 provides a detailed overview of all abbreviations used.

**Figure 3.**
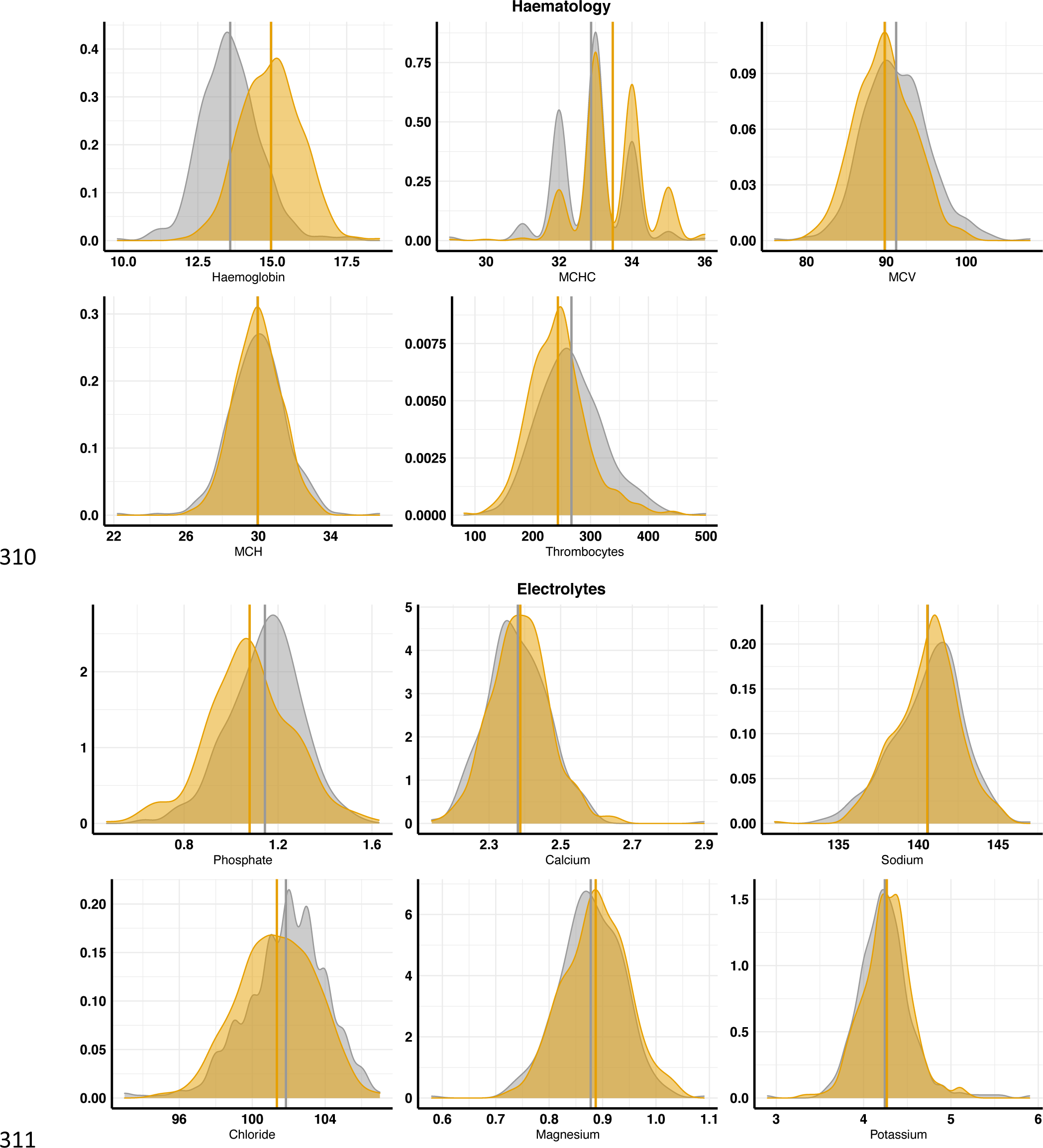
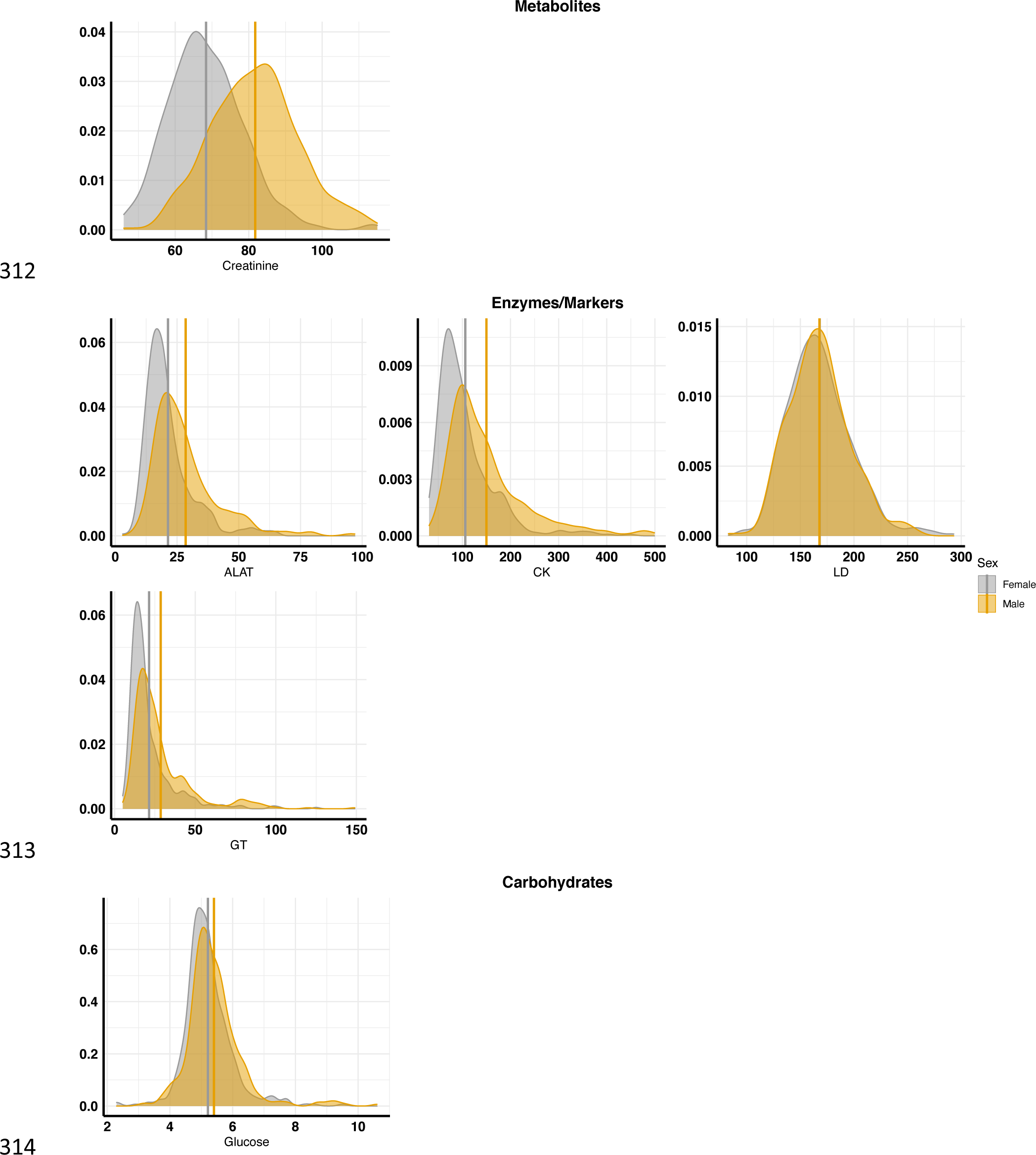
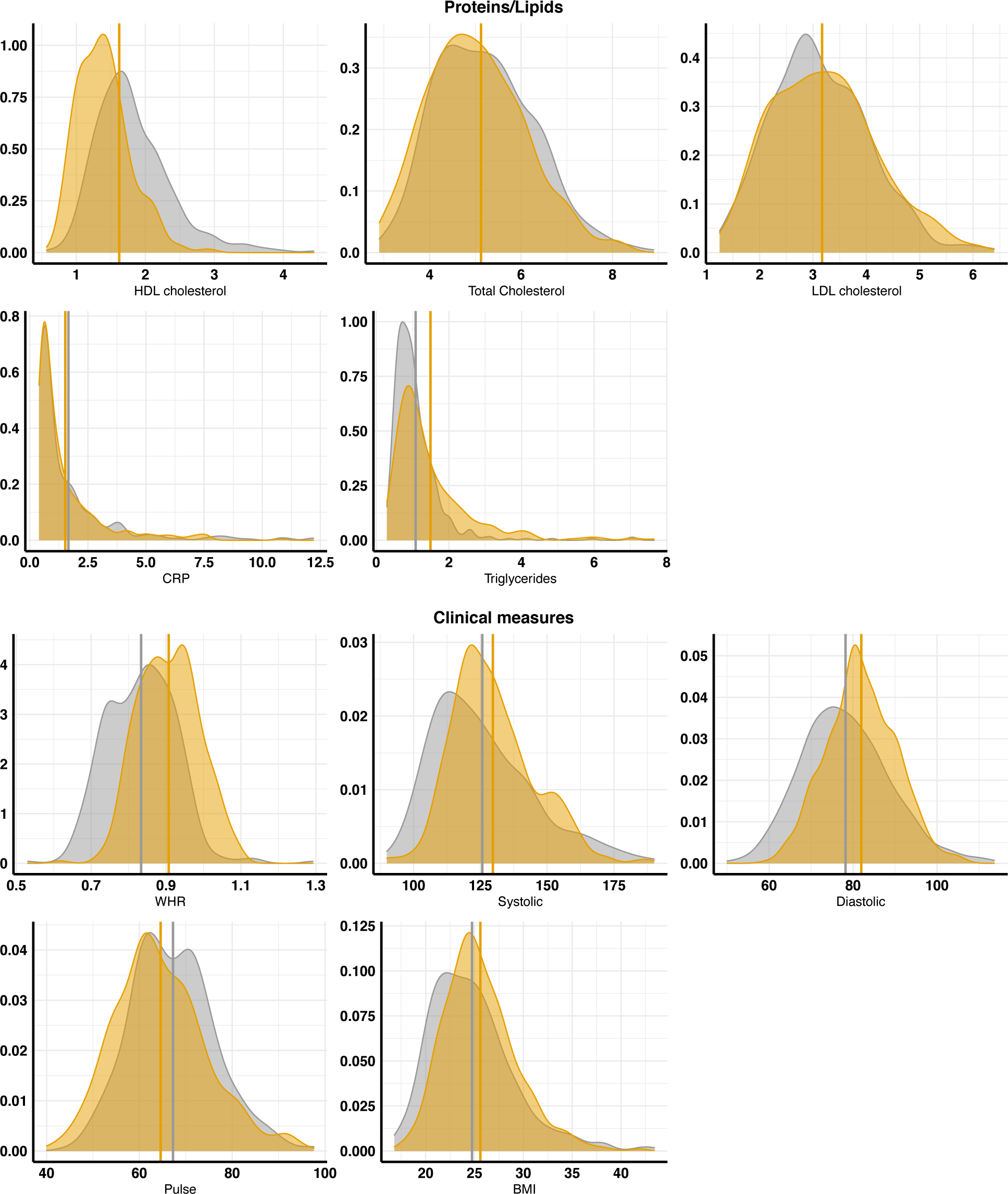
Distribution of the cardiometabolic risk factors. Density plots for each variable, split by sex (male = orange, female = grey). Vertical lines represent mean values for each sex. See SI Table 2 for reference (normal/healthy) range for each variable.

**Figure 4.**
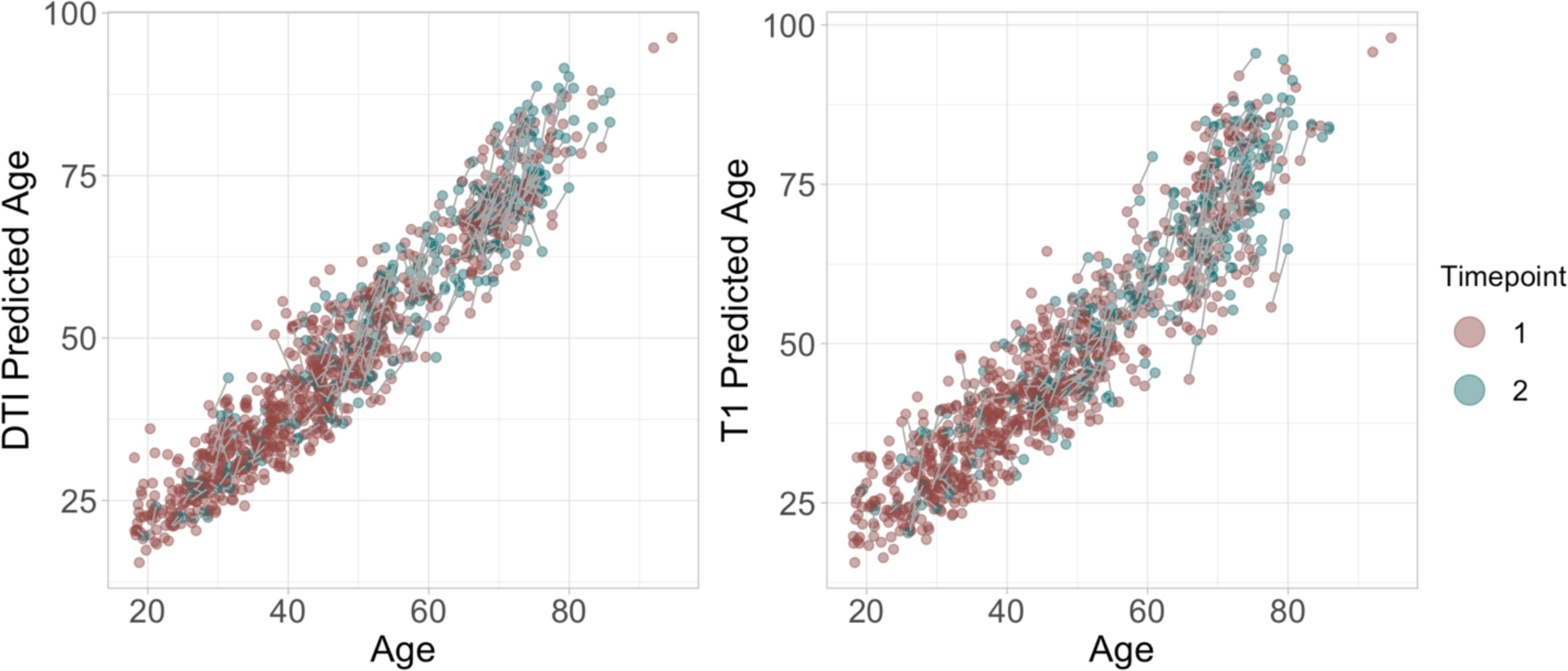
Predicted age as a function of age. The figure shows baseline brain age in red and follow up brain age in green.

**Figure 5.**
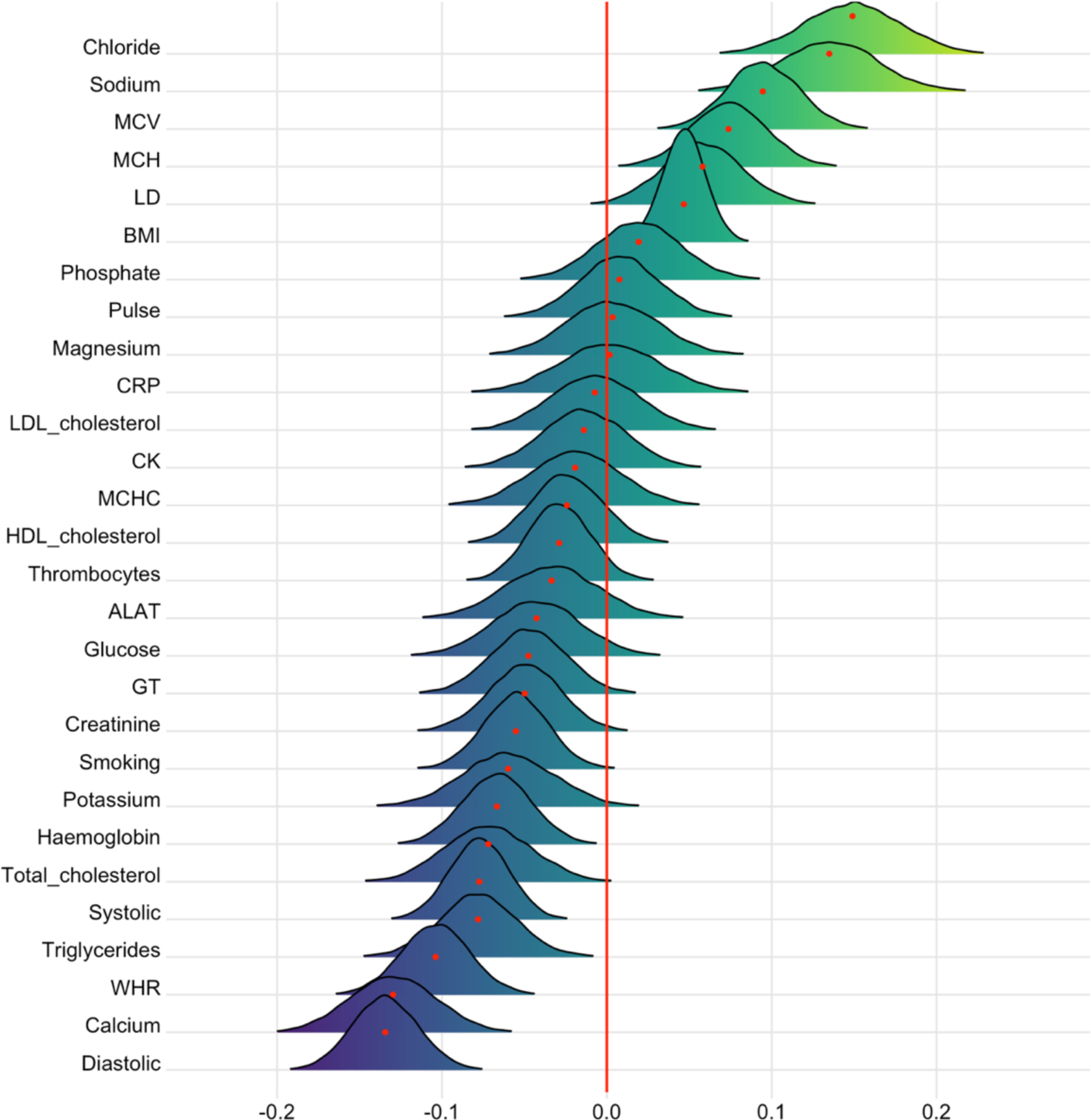
Associations between cardiometabolic risk factors and time. The figure shows posterior distributions of the estimates of the coefficient. Estimates for time on each variable with red dot in each plot representing mean value. Colour scale follows direction evidence. Width of distribution represents the uncertainty of the parameter estimates.

**Figure 6.**
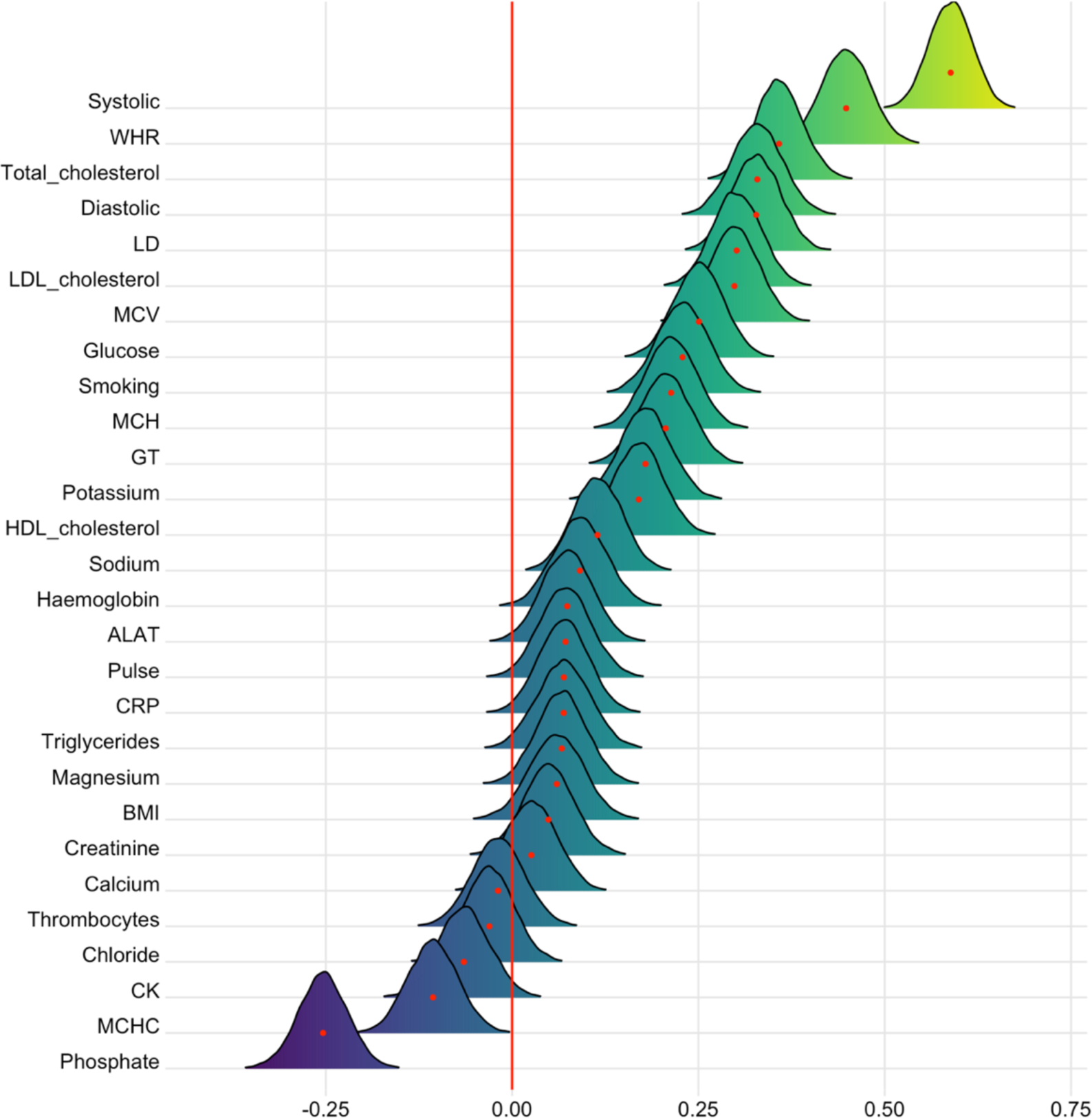
Associations between cardiometabolic risk factors and age. The figure shows posterior distributions of the estimates of the coefficient. Estimates for age on each variable.

**Figure 7.**
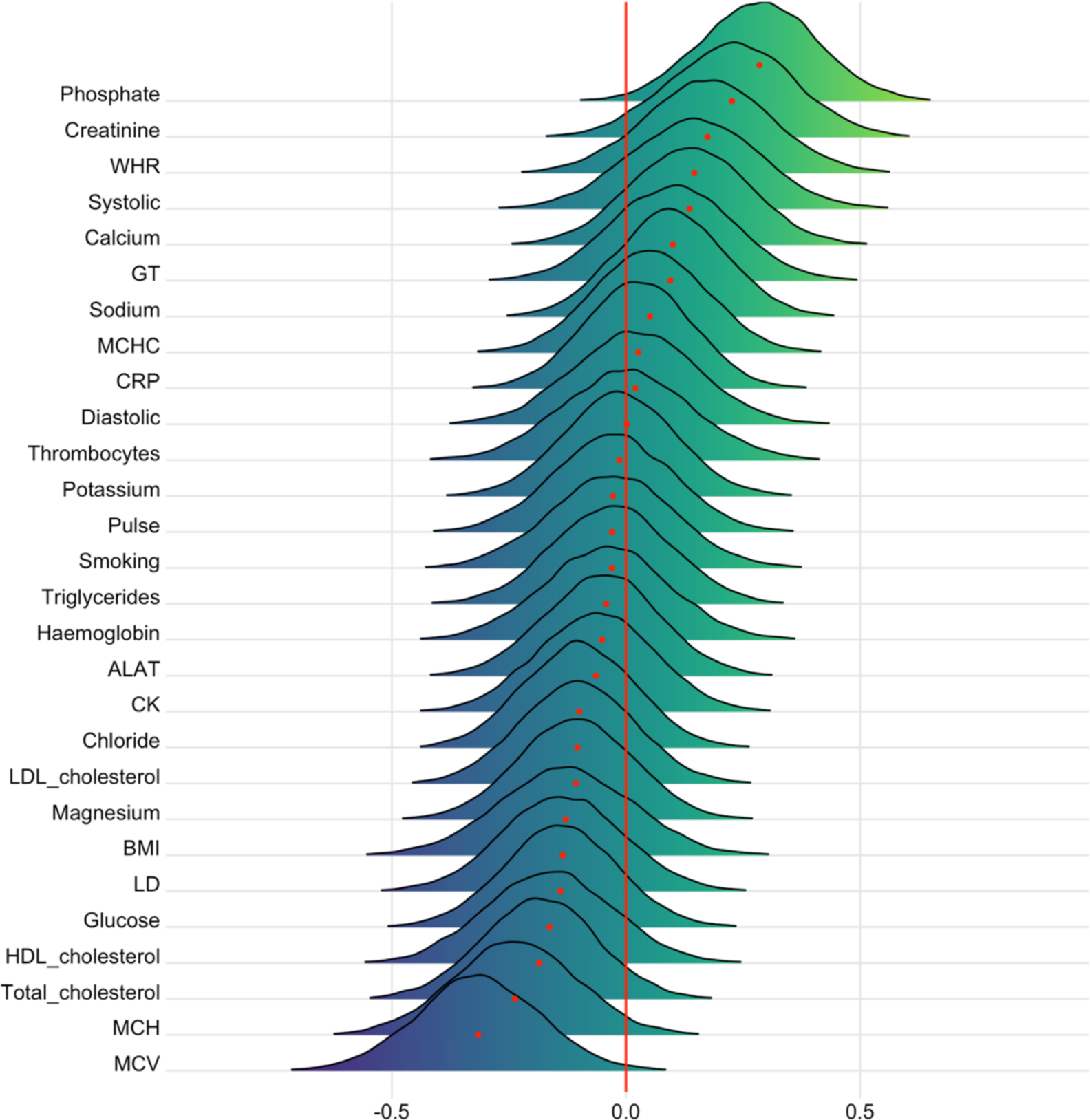
Associations between cardiometabolic risk factors and DTI BAG. The figure shows posterior distributions of the estimates of the coefficient. Estimates for each variable on DTI BAG.

**Figure 8.**
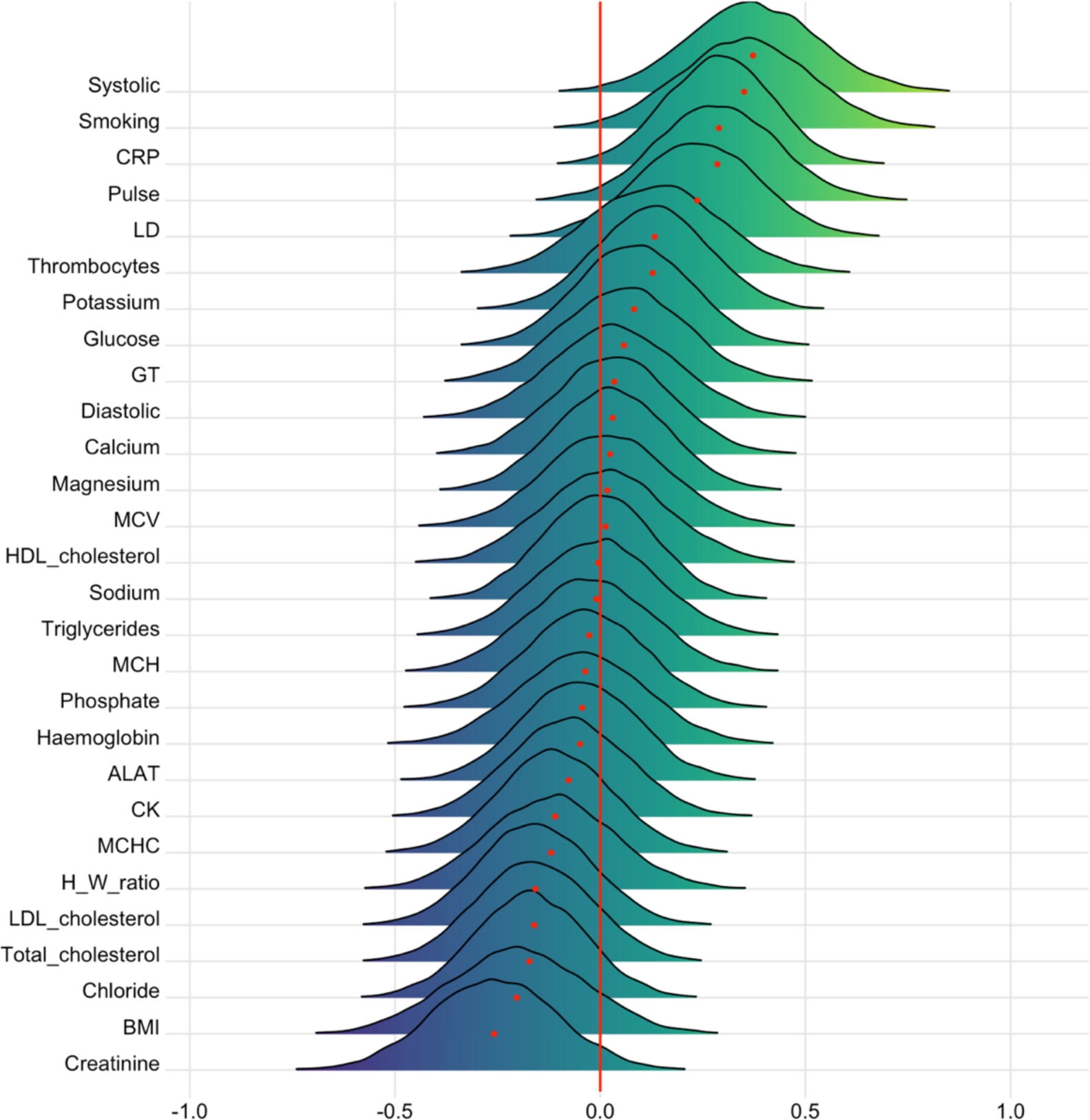
Associations between cardiometabolic risk factors and T1 BAG. The figure shows posterior distributions of the estimates of the coefficient. Estimates for each variable on T1 BAG.

To visualise the associations between the CMRs, hierarchical clustering of the variables was performed using ‘hclust’, part of the ‘stats’ package in R (R Core Team, 2012), which uses the complete linkage method to form clusters. Five cluster groups were revealed. Figure 2 provides a visualisation of the hierarchical clustering, including the full (upper diagonal) and partial (lower diagonal) correlations.

### 2.8. Statistical analysis

All statistical analyses were carried out using R, version 3.6.0 (www.r-project.org/) (R Core Team, 2012). To investigate the associations between the CMRs and BAG, we carried out Bayesian multilevel models in ‘Stan’ (Stan Development Team, 2019) using the *brms* (Bürkner, 2017, 2018) package in R (R Core Team, 2012). For descriptive purposes, we first tested associations between BAG and time. Here, BAG (for T1 and DTI separately) was entered as the dependent variable while timepoint was entered as the independent variable. Second, we tested associations between each CMR and time and age. Here, timepoint and age were entered as the independent variable (in separate analyses). Thirdly, to address the primary aim of the study, we tested for associations between brain age gap and each CMR across time. Here, BAG (for T1 and DTI separately) was entered as the dependent variable with each CMR separately entered as the independent fixed effects variable along with age, sex, and time, with subject ID as random effects. Fourth, in order to test our hypothesis that the associations between cardiometabolic risk and BAG vary as a function of age both cross-sectionally and longitudinally, interaction effects of CMR and age on BAG, and CMR and time on BAG, were included in the models as additional fixed effects. For each model, timepoint and age were included in the models where appropriate, while sex was added to every model. In order to prevent false positives and to regularise the estimated associations, we defined a strong normal prior around zero with a standard deviation of 0.3 for all coefficients bar BAG ∼ time. For each coefficient of interest, we report the mean estimated value and its uncertainty measured by the 95% credible interval of the posterior distribution. We calculated Bayes factors (BF) using the Savage-Dickey method (Wagenmakers et al., 2010). For a pragmatic guide on Bayes factor (BF) interpretation, see SI Table 3.

## 3. Results

### 3.1. Brain age prediction

Within the training sample, the correlation between predicted and chronological age was *r* = 0.91 95% CI [0.89, 0.92] for the DTI model, and *r* = 0.90 [0.87, 0.92] for the model based on T1-weighted data. Applying the model to the test sample resulted in a correlation between predicted and chronological age of *r* = 0.85 [0.83, 0.87] for the DTI model, and *r* = 0.85 [0.84, 0.87] for the model based on T1-weighted data. SI Figure 9 shows the correlations before and after age-bias correction. R^2^, RMSE, and MAE are provided in Table 2.

**Figure 9.**
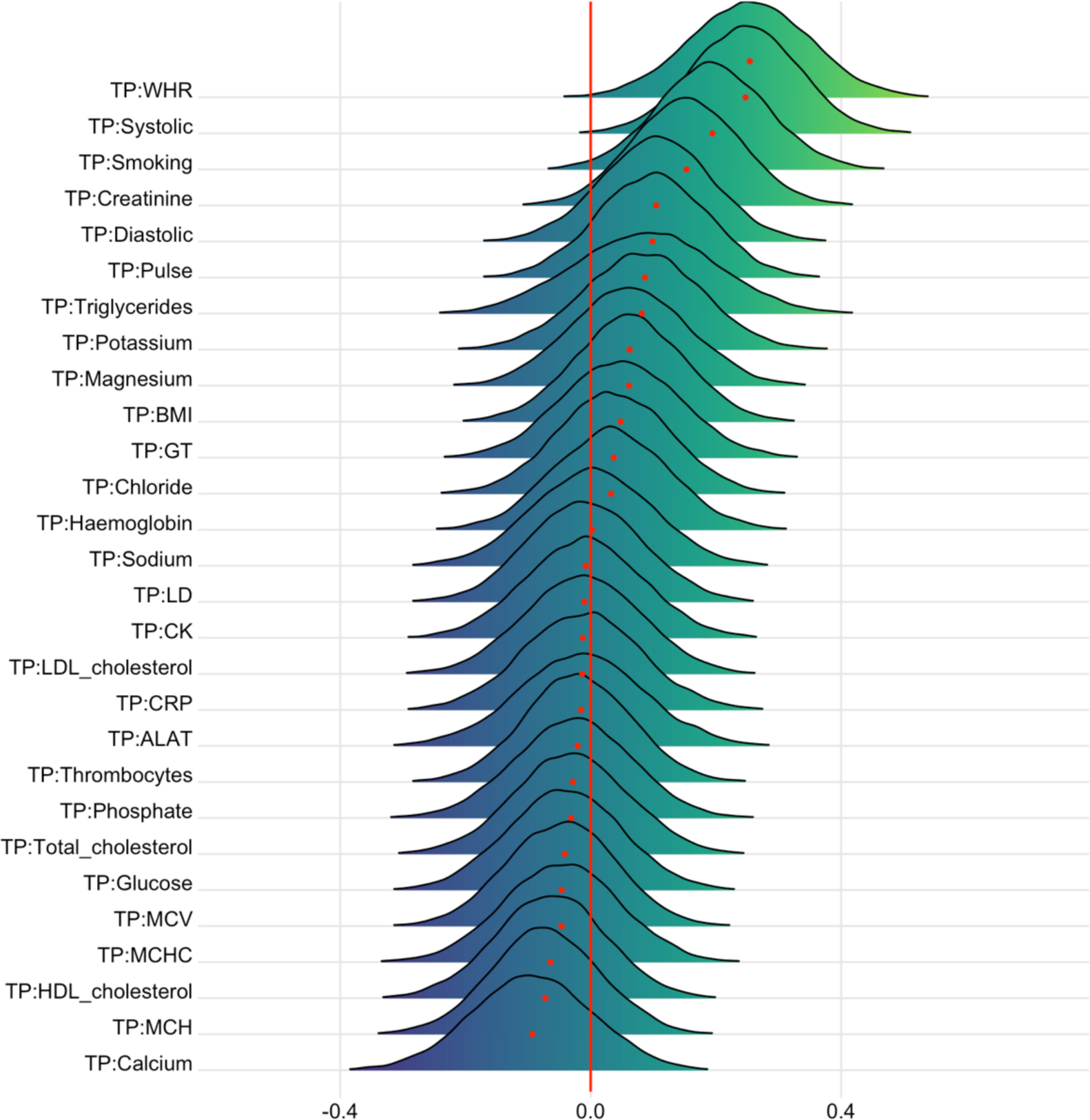
Interaction effects between cardiometabolic risk factors and time on DTI BAG. The figure shows posterior distributions of the estimates of the coefficient. Estimates for the interaction effect of time and each CMRs on DTI BAG.

### 3.2. Cardiometabolic risk factors

#### 3.2.1. **Descriptive statistics**

**Table 3.**
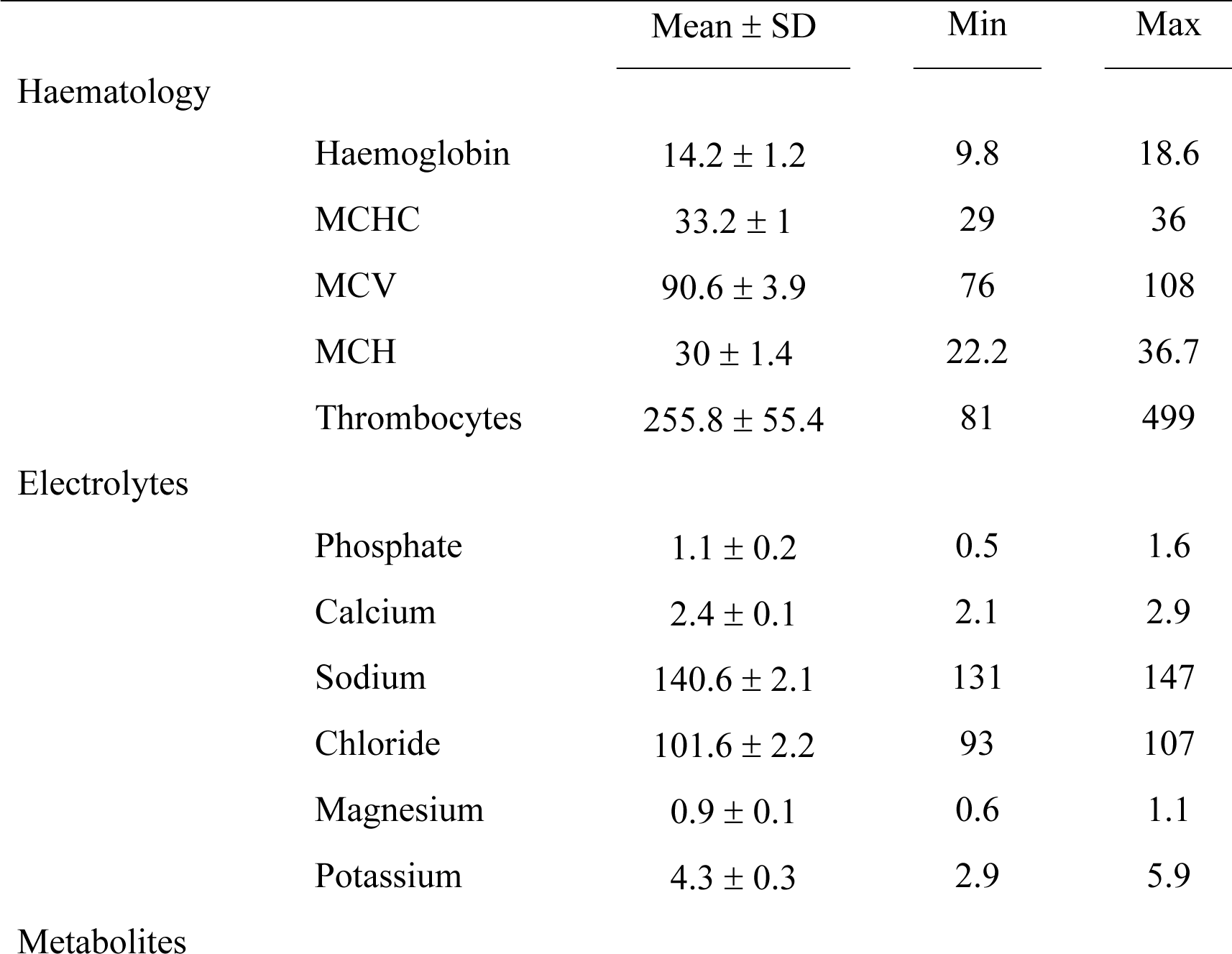
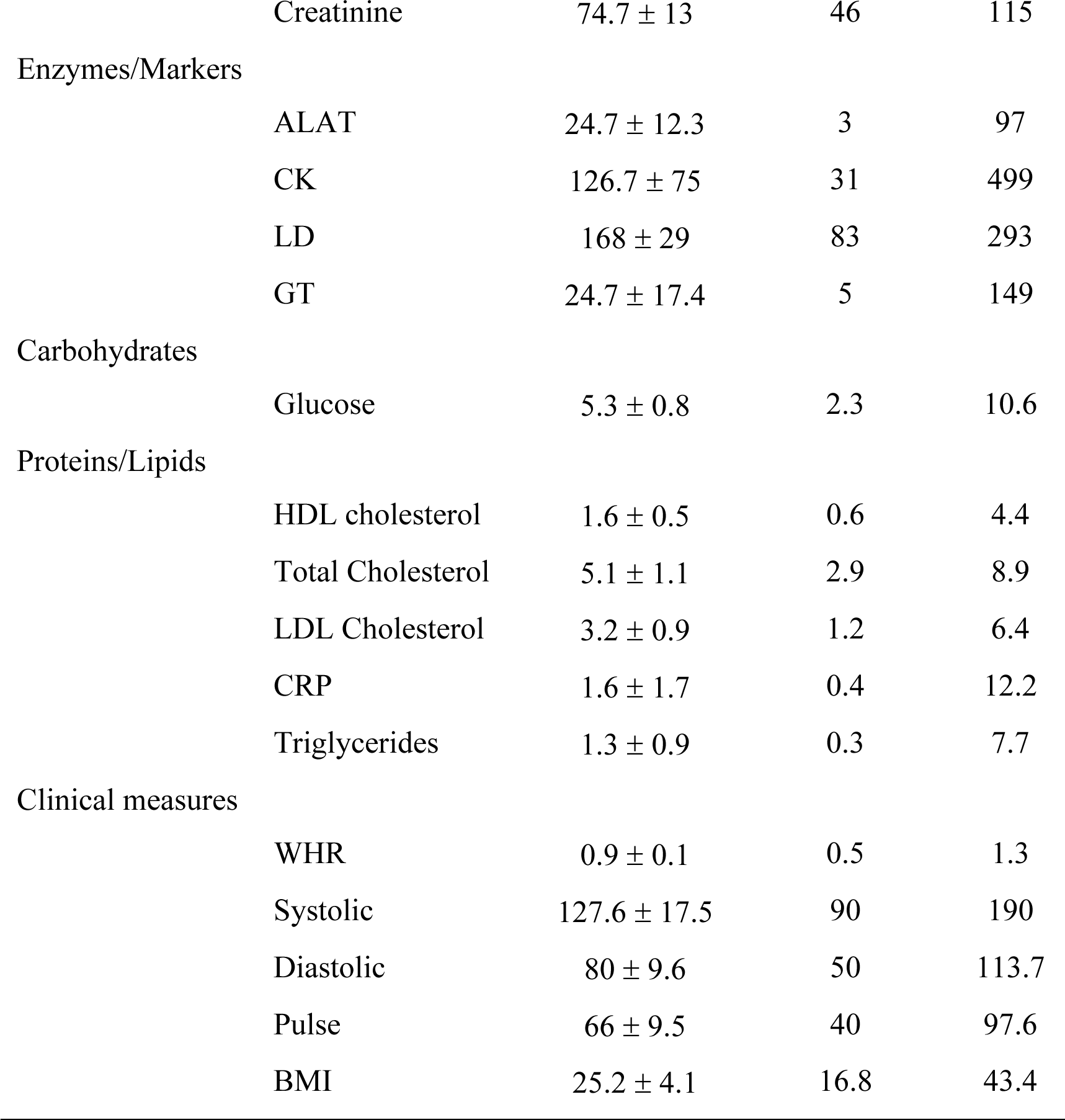
Descriptive statistics at baseline for each variable bar smoking, which is summarised in its own table due to its ordinal nature.

|  |  | Mean ± SD | Min | Max |
| --- | --- | --- | --- | --- |
| Haematology |  |  |  |  |
|  | Haemoglobin | 14.2 ± 1.2 | 9.8 | 18.6 |
|  | MCHC | 33.2 ± 1 | 29 | 36 |
|  | MCV | 90.6 ± 3.9 | 76 | 108 |
|  | MCH | 30 ± 1.4 | 22.2 | 36.7 |
|  | Thrombocytes | 255.8 ± 55.4 | 81 | 499 |
| Electrolytes |  |  |  |  |
|  | Phosphate | 1.1 ± 0.2 | 0.5 | 1.6 |
|  | Calcium | 2.4 ± 0.1 | 2.1 | 2.9 |
|  | Sodium | 140.6 ± 2.1 | 131 | 147 |
|  | Chloride | 101.6 ± 2.2 | 93 | 107 |
|  | Magnesium | 0.9 ± 0.1 | 0.6 | 1.1 |
|  | Potassium | 4.3 ± 0.3 | 2.9 | 5.9 |
| Metabolites |  |  |  |  |
|  | Creatinine | 74.7 ± 13 | 46 | 115 |
| Enzymes/Markers |  |  |  |  |
|  | ALAT | 24.7 ± 12.3 | 3 | 97 |
|  | CK | 126.7 ± 75 | 31 | 499 |
|  | LD | 168 ± 29 | 83 | 293 |
|  | GT | 24.7 ± 17.4 | 5 | 149 |
| Carbohydrates |  |  |  |  |
|  | Glucose | 5.3 ± 0.8 | 2.3 | 10.6 |
| Proteins/Lipids |  |  |  |  |
|  | HDL cholesterol | 1.6 ± 0.5 | 0.6 | 4.4 |
|  | Total Cholesterol | 5.1 ± 1.1 | 2.9 | 8.9 |
|  | LDL Cholesterol | 3.2 ± 0.9 | 1.2 | 6.4 |
|  | CRP | 1.6 ± 1.7 | 0.4 | 12.2 |
|  | Triglycerides | 1.3 ± 0.9 | 0.3 | 7.7 |
| Clinical measures |  |  |  |  |
|  | WHR | 0.9 ± 0.1 | 0.5 | 1.3 |
|  | Systolic | 127.6 ± 17.5 | 90 | 190 |
|  | Diastolic | 80 ± 9.6 | 50 | 113.7 |
|  | Pulse | 66 ± 9.5 | 40 | 97.6 |
|  | BMI | 25.2 ± 4.1 | 16.8 | 43.4 |

**Table 4.** Smoking at baseline.

|  | Frequency (%) |
| --- | --- |
| Never smoked | 593 (75.1) |
| Previous smoker | 127 (16.) |
| Current smoker | 70 (8.9) |

### 3.3. Bayesian multilevel models

#### 3.3.1. **Effects of time on brain age gaps**

Figure 4 shows predicted age for each model plotted as a function of age. Bayesian modelling revealed higher DTI and T1 based BAG at follow-up than baseline (SI Figure 10).

**Figure 10.**
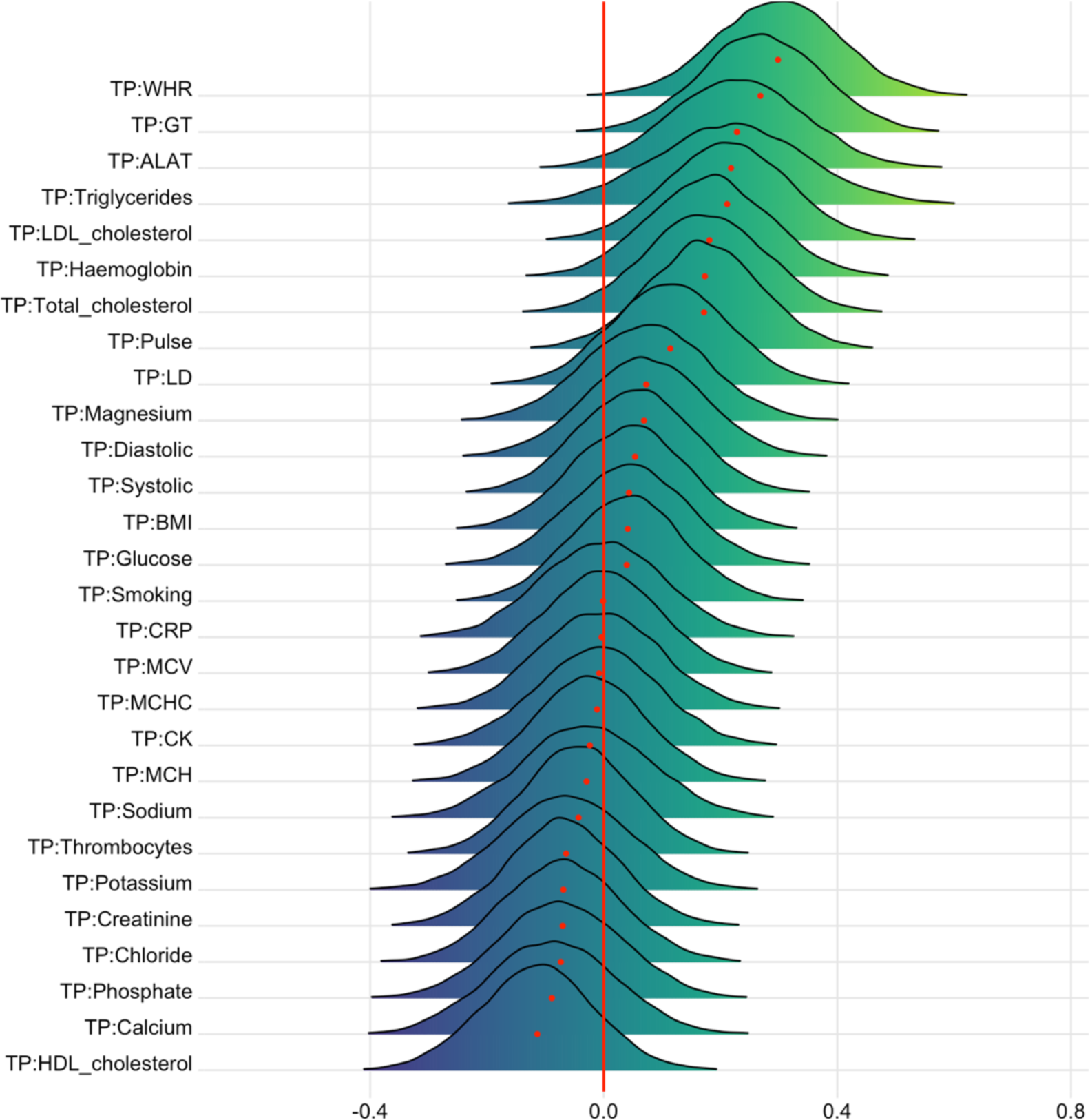
Interaction effects between cardiometabolic risk factors and time on T1 BAG. The figure shows posterior distributions of the estimates of the interaction effect of time and each variable on T1 BAG. Width of the distribution represents the uncertainty of the parameter estimates.

#### 3.3.2. Effects of time and age on CMRs

Figure 5 shows the posterior distributions for estimates of the coefficient for time on each variable. Full table of results for time and age effects on each variable can be seen in SI Table 4. Supplementary visualisation of the effects of time and age on a selection of the CMRs can be seen in SI Figures 11, 12, and 13.

**Figure 11.**
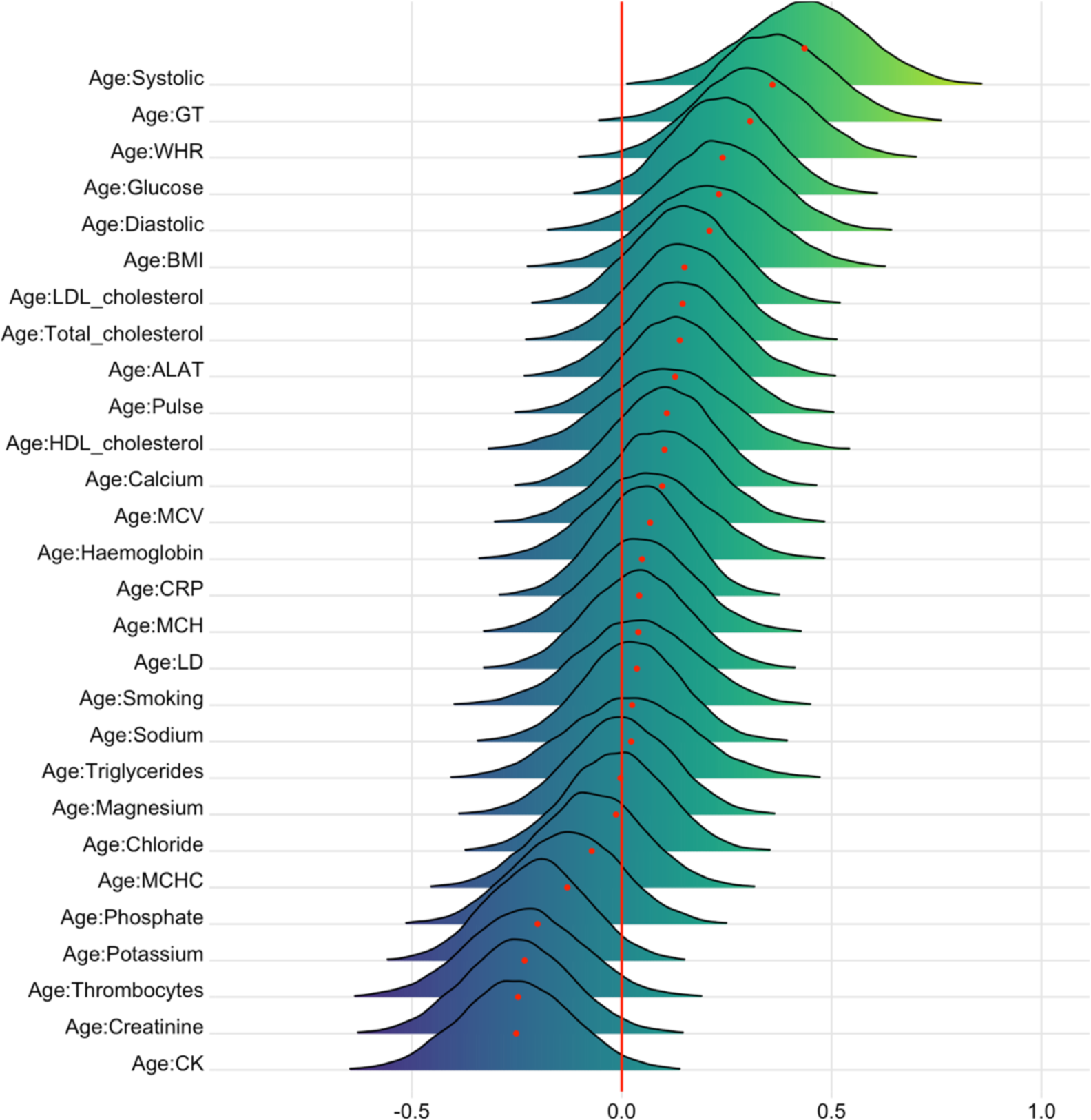
Interaction effects between cardiometabolic risk factors and age on DTI BAG. The figure shows posterior distributions of the estimates for the interaction effect between age and each variable on DTI BAG, with red dot representing mean value. Width of the distribution represents the uncertainty of the parameter estimates.

**Figure 12.**
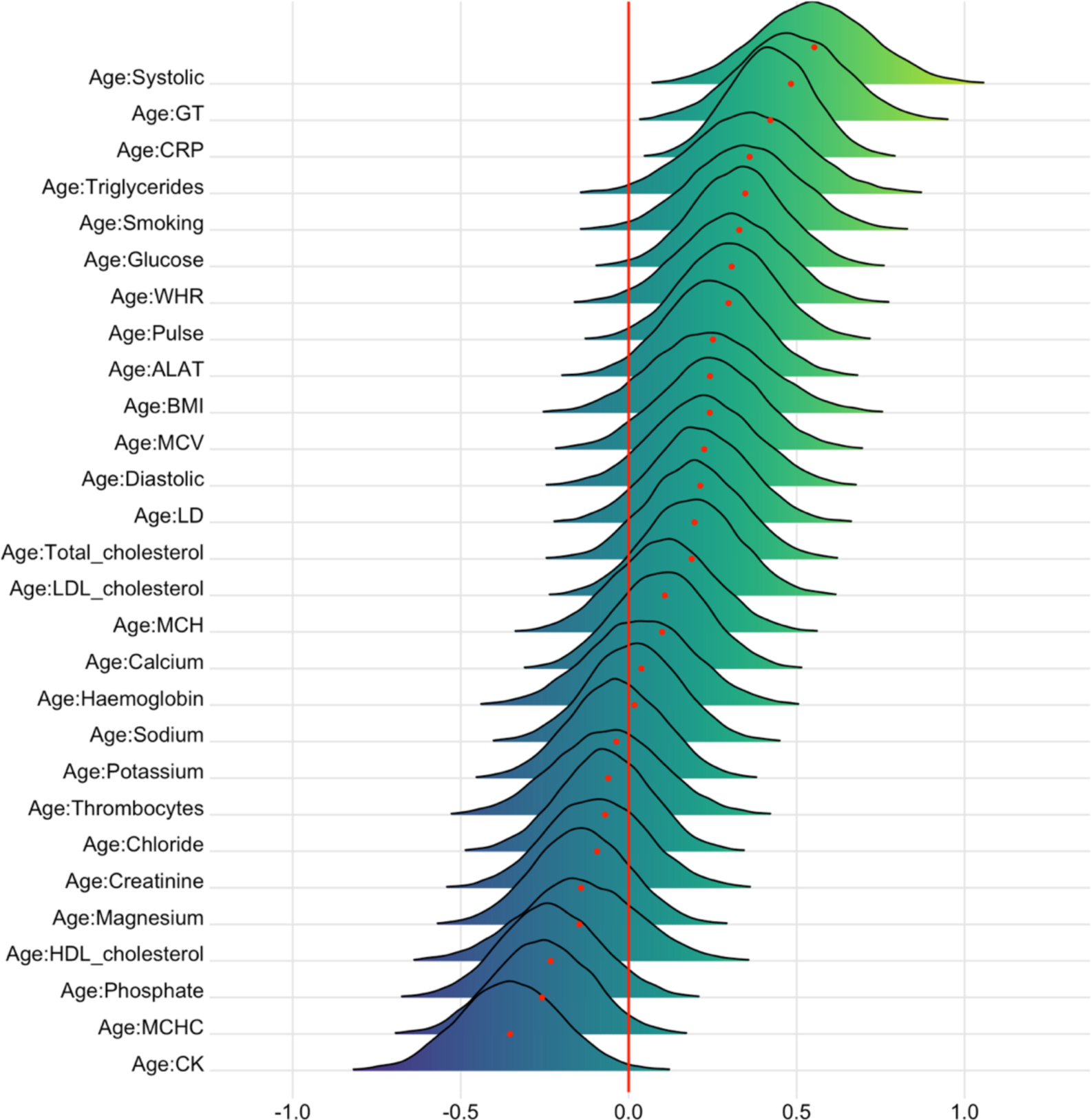
Interaction effects between cardiometabolic risk factors and age on T1 BAG. The figure shows posterior distributions of the estimates for the interaction effect between age and each variable on T1 BAG, with red dot representing mean value. Width of the distribution represents the uncertainty of the parameter estimate.

Briefly, the tests confirmed extreme evidence (BF < 0.01) in favour of an association between time and calcium (*β* = -0.13), WHR (*β* = -0.10), sodium (*β =* 0.13), chloride (*β =* 0.15), MCV (*β =* 0.10), systolic blood pressure (*β =* -0.08), and diastolic blood pressure (*β =* -0.13). Very strong evidence was provided for BMI (BF = 0.03, *β =* 0.05), while strong evidence was provided for triglycerides (BF = 0.08, *β =* -0.08), haemoglobin (BF = 0.07, *β =* -0.07), and MCH (BF = 0.08, *β =* 0.08). Strong evidence was also provided in favour of no (null) association between time and pulse (BF = 11.8, *β =* 0.008), magnesium (BF = 10.8, *β =* 0.003), and LDL cholesterol (BF = 10.7, *β =* -0.007).

Figure 6 shows the posterior distributions for estimates of the coefficient for age on each variable. The tests revealed extreme evidence (BF < 0.01) in favour of an age association for phosphate (*β =* -0.25), HDL cholesterol (*β =* 0.17), glucose (*β =* 0.25), GT (*β =* 0.21), WHR (*β =* 0.45), LD (*β =* 0.33), MCV (*β =* 0.30), MCH (*β =* 0.21), total cholesterol (*β =* 0.36), LDL cholesterol (*β =* 0.30), systolic (*β =* 0.59) and diastolic blood pressure (*β =* 0.33), potassium (*β =* 0.18), and smoking (*β =* 0.23). The models revealed moderate evidence in favour of no changes over time for thrombocytes (BF = 7.51, *β =* -0.02), calcium (BF = 6.68, *β =* 0.03), creatinine (BF = 3.24, *β =* 0.05), and chloride (BF = 5.82, *β =* -0.03).

#### 3.3.3. Associations between BAG and CMRs

Figures 7 and 8 show posterior distributions of the estimates of the coefficient reflecting the associations between each CMR and BAGs, and SI Table 5 and 6 show full table of results. Credible intervals and evidence ratios can be found in SI Figure 14. The tests revealed moderate evidence in favour of an association between DTI BAG and phosphate (BF = 0.17, *β =* 0.29) and MCV (BF = 0.14, *β =* -0.32), and anecdotal evidence for WHR (BF = 0.89, *β =* 0.17), creatinine (BF = 0.6, *β =* 0.23), MCH (BF = 0.46, *β =* -0.24), and total cholesterol (BF = 0.76, *β =* -0.19).

Moderate evidence in favour of an association with T1 BAG was provided for systolic blood pressure (BF = 0.13, *β =* 0.37), smoking (BF = 0.17, *β =* 0.35), pulse (BF = 0.3, *β =* 0.29), and CRP (BF = 0.21, *β =* 0.29), and anecdotal evidence for BMI (BF = 0.86, *β =* -0.20), LD (BF = 0.55, *β =* 0.24), and creatinine (BF = 0.52, *β =* -0.26). No results provided moderate or stronger evidence (BF>3) in favour of the null.

#### 3.3.4. Interaction effects of time and CMRs on brain age gap

Figures 9 and 10 show posterior distributions reflecting estimates of the coefficient for the interaction between time and each CMRs on DTI and T1 BAGs. SI Table 7 and 8 show full table of results. Credible intervals and evidence ratios can be found in SI Figure 15. For DTI BAG, the evidence supporting an interaction with time was strong for WHR (BF = 0.09, *β =* 0.25) and systolic blood pressure (BF = 0.07, *β =* 0.25), indicating faster pace of brain ageing among people with higher WHR and systolic blood pressure. This is visualised in SI Figure 16. The models further indicated moderate evidence for smoking (BF = 0.31, *β =* 0.19) and anecdotal evidence for creatinine (BF = 0.73, *β =* 0.15).

For T1 BAG, the evidence supporting an interaction with time was strong for WHR (BF = 0.06, *β =* 0.30), indicating faster pace of brain ageing among people with higher WHR. The models also revealed moderate evidence for GT (BF = 0.11, *β =* 0.27), and anecdotal for pulse (BF = 0.63, *β =* 0.17), triglycerides (BF = 0.58, *β =* 0.22), ALAT (BF = 0.37, *β =* 0.23), haemoglobin (BF = 0.64, *β =* 0.18), total cholesterol (BF = 0.72, *β =* 0.17), and LDL cholesterol (BF = 0.38, *β =* 0.21).

Thrombocytes (BF = 3.34, *β =* -0.02), CRP (BF = 3.18, *β =* -0.01), phosphate (BF = 3.11, *β =* -0.03), ALAT (BF = 3.06, *β =* -0.02), CK (BF = 3.23, *β =* -0.01), LD (BF = 3.2, *β = -*0.01), sodium (BF = 3.1, *β <* 0.01), chloride (BF = 3.16, *β =* 0.04), total cholesterol (BF = 3.14, *β =* - 0.03), and LDL cholesterol (BF = 3.19, *β =* -0.01) showed moderate evidence in favour of no interaction effect with time on DTI BAG (SI Table 3). For T1 BAG, only MCV showed moderate evidence in favour of no association (BF = 3, *β <* 0.01).

#### 3.3.5. Interaction effects of age and CMRs on changes in brain age gap

Figures 11 and 12 show posterior distributions of the estimates of the coefficient for the interaction between age and each CMR on DTI and T1 BAGs. See SI Table 9 and 10 for full table of results. Credible intervals and evidence ratios can be found in SI Figure 17. The analysis provided strong support of an interaction effect with age on DTI BAG for GT (BF = 0.09, *β =* 0.36) and systolic blood pressure (BF = 0.02, *β =* 0.44), indicating that GT and systolic blood pressure are more important predictors of brain age with increasing age. This is visualised in SI Figure 18. The models further indicated moderate support for WHR (BF = 0.18, *β =* 0.31), and anecdotal support for thrombocytes (BF = 0.56, *β =* -0.23), glucose (BF = 0.36, *β =* 0.24), BMI (BF = 0.72, *β =* 0.21), CK (BF = 0.38, *β = -*0.25), creatinine (BF = 0.4, *β = -*0.25), diastolic blood pressure (BF = 0.51, *β =* 0.23), and potassium (BF = 0.6, *β = -*0.20).

The support of an interaction effect with age on T1 BAG was strong for CRP (BF = 0.01, *β =* 0.42) and systolic blood pressure (BF = 0.01, *β =* 0.55), indicating that CRP and systolic blood pressure are increasingly important predictors of BAG with increasing age. The models further indicated moderate evidence for pulse (BF = 0.25, *β =* 0.30), glucose (BF = 0.15, *β =* 0.33), triglycerides (BF = 0.19, *β =* 0.36), WHR (BF = 0.3, *β =* 0.31), CK (BF = 0.13, *β = -*0.35), and smoking (BF = 0.18, *β =* 0.35), and anecdotal for phosphate (BF = 0.63, *β = -*0.23), BMI (BF = 0.67, *β =* 0.24), ALAT (BF = 0.44, *β =* 0.25), LD (BF = 0.72, *β =* 0.21), MCHC (BF = 0.41, *β = -*0.26), MCV (BF = 0.62, *β =* 0.24), total cholesterol (BF = 0.88, *β =* 0.20), and LDL cholesterol (BF = 0.88, *β =* 0.19). No results provided moderate or stronger evidence (BF>3) in favour of the null hypothesis.

## 4. Discussion

Brain and cognitive ageing is highly heterogenous and may involve a range of biological processes. Cardiometabolic risk factors are associated with increased risk of brain disorders, and a better understanding of the links between brain ageing and malleable indicators of cardiometabolic health may provide a window of opportunity for interventions. The current cross-sectional and longitudinal findings support that higher cardiometabolic risk is associated with faster brain ageing. Both the overall brain age gap and the rates of change were associated with a range of CMRs, including anthropomorphic measures, blood lipids, lifestyle factors (smoking), and blood pressure.

### 4.1. Associations between CMRs and age, and interactions with age

Age showed credible associations with several CMRs, including phosphate, HDL cholesterol, glucose, GT, WHR, LD, MCV, MCH, total cholesterol, LDL cholesterol, systolic and diastolic blood pressure, potassium, and smoking. Interaction effects for age and CMRs on DTI BAG were evident for GT, systolic blood pressure, and WHR. For T1 BAG, age interaction effects were evident for CRP and systolic blood pressure, pulse, triglycerides, WHR, CK, and smoking.

In general, these associations are in line with previous studies showing associations between CMR and age-related neurodegenerative diseases and cognitive decline. For example, higher serum phosphate, an element filtered by the kidney, is associated with increased risk of incident dementia (Li et al., 2017), and risk of brain haemorrhage (Yamada et al., 2016), while low serum phosphate level is associated with cerebral β-amyloid deposition (Park et al., 2017), increased risk of brain infarction in haemodialysis patients (Yamada et al., 2016), and lower composite score in relation to cognitive function (Basheer et al., 2016).

Previous research has also found that high serum potassium levels were associated with mild cognitive impairment (Vintimilla et al., 2018), while high glucose levels were associated with low gray matter density and FA (Weinstein et al., 2015). Higher GT levels, as an index of liver function, has previously been associated with brain volume shrinkage in patients with alcohol dependence (Chen et al., 2012), brain infarcts in a healthy population (Nam et al., 2019), and cardiovascular mortality (Ruttmann et al., 2005).

### 4.2. Associations between CMR and brain ageing, and interactions with time

Supporting the hypothesized link between cardiometabolic risk and brain ageing, our findings demonstrated associations between several CMRs and BAG. Strongest evidence was found for phosphate and MCV for DTI BAG, and systolic blood pressure, smoking, pulse, and CRP for T1 BAG, indicating older-appearing brains in people with poorer cardiometabolic health. Further, our longitudinal analyses revealed that the rate of brain ageing across the study period was influenced by cardiometabolic risk, with strong evidence for WHR for both BAG models, and systolic blood pressure for DTI BAG. In addition, moderate evidence of smoking was found for DTI BAG. For these effects, reduced cardiometabolic health was associated with increased rate of brain ageing.

In general, these associations are in line with previous studies showing associations between CMRs and age-related cognitive decline, with higher MCV levels being associated with reduced episodic memory, affected global cognitive function and mental status (Chen et al., 2020; Gamaldo et al., 2014), in addition to an increased risk of cerebrovascular and cardiovascular related deaths (Wu, 2018). Moreover, related health markers of red blood cell measures (MCHC) have previously been associated with higher depressive symptom scores (Lee et al., 2017).

Higher levels of CRP, a systemic marker of inflammation, have previously been associated with smaller temporal lobes (Bettcher et al., 2012), reduced working memory, smaller cortical thickness in frontal, insula, and temporal brain regions (Jacomb et al., 2018), worse performance in tests assessing executive functions, reduced global FA (Wersching et al., 2010), and increased cerebral myoinositol (Eagan et al., 2012). Although the mechanisms remain unclear, elevated CRP has also been found in patients with acute psychosis and schizophrenia (Jacomb et al., 2018).

While no direct support from studies looking at WHR and brain age gap currently exists, previous studies have reported associations between obesity and white matter DTI (FA and MD), white matter volume (Karlsson et al., 2013) and brain age (Kolenic et al., 2018; Ronan et al., 2016). Additionally, research investigating the association between adipose tissue and brain health has recently revealed significant negative associations between BMI and white matter surface area and cortical gray matter volume, and between WHR and caudate volume (Gurholt et al., 2020).

Our results demonstrating that elevated systolic blood pressure and smoking were associated with faster brain ageing over time is in line with previous cross-sectional studies, with systolic blood pressure reportedly being associated with white matter BAG (de Lange et al., 2020) and reduced cerebral vascular density (Williamson et al., 2018). Similarly, smoking has also been associated with decreased total brain volume (Reiman et al., 2008) and reduced cerebral vascular density (Williamson et al., 2018). Moreover, longitudinal studies have reported higher rates of annual white matter lesion progression in subjects with increased systolic blood pressure (Verhaaren et al., 2013).

Albeit with moderate evidence, the rate of brain ageing was also associated with increased pulse and several key blood biomarkers reflecting various aspects of cardiometabolic health, including creatinine, GT, triglycerides, ALAT, total and LDL cholesterol, and haemoglobin. These findings jointly contribute to the larger picture of modifiable CMRs influencing brain ageing. Additionally, the findings are largely in line with previous studies demonstrating more white matter hyperintensities in people with lower haemoglobin levels, and less coherent white matter in people with high total and LDL cholesterol and triglyceride levels (Williams et al., 2013).

### 4.3. Future research and recommendations for treatment

In common with other imaging based surrogate markers, brain predicted age should be understood as not only a phenomenon impacted by the effects of ageing, but also by the effects of a lifetime of exposure to positive and negative lifestyles and environments, coupled with a genetic component that plays a crucial role in the variation. Despite not being able to disentangle the contribution of each of these components when looking at an individual’s predicted age and subsequent brain age gap, this disparity between chronological and predicted age still provides us with an individualised marker of deviation from the expected value. And with this, recommendations for treatment can focus on the individual and target management of risk.

Alternatively, early intervention strategies that place their focus on prevention rather than management of risk may be more beneficial. For example, Williamson et al. (2018) found that cardiovascular health in early adulthood relates to brain atrophy in later life. McEvoy et al. (2015) found that white matter alterations appear early in the course of hypertension and may persist despite adequate treatment. The implication of this suggests that preventive strategies that promote cardiometabolic health early may be more beneficial in prolonging healthy brain ageing. Moreover, maintaining the structure of the brain in a younger state may have profound effects on delaying the onset of age-related neurodegenerative diseases (Qiu & Fratiglioni, 2015; Steffener, 2016). Informed by studies showing that higher levels of physical exercise (Steffener, 2016) and meditation (Luders et al., 2016) are associated with lower brain ageing, interacting with modifiable CMRs may change risk trajectories and prevent progression to disease for a manifold of cardiovascular and neurodegenerative diseases as well as mental disorders (Ringen et al., 2014, 2018; Schmitt et al., 2018).

## 4.4. Strengths and limitations

The current study had several strengths. As there is both variability in the brain ageing process from person to person (Aycheh et al., 2018), and variability in CMRs, the current study benefitted from a mixed cross-sectional and longitudinal design, whereby changes can be tracked across timepoints. For brain age, the prediction models had high accuracy, and separate diffusion and T1-weighted brain age gaps provided insight into modality-specific impact of CMRs. Most brain-age models use only T1-weighted structural MRI but changes in white matter microstructure and coherence may precede alterations that may not be detected by T1-weighted MRI (Cole, 2020).

Some limitations must also be addressed. The sample is predominantly ethnic Northern European/Scandinavian, restricting our ability to generalise to the wider public and other population groups of generally higher or lower risk than our sample. Moreover, the sample is generally healthy, and biases due to non-random attrition could be introduced. Contrarily, the range of the values for many blood test and pressure measurements reveal incidental indication of possible kidney failure, anaemia, platelet disorder, hyperlipidaemia, and hypertension. Future studies should utilise a more comprehensive cardiometabolic risk assessment detailing dietary routines, alcohol intake, psychosocial stress, and physical activity, which jointly have been shown to account for 90% of the population-attributable risk of myocardial infarction in men and 94% in women (Yusuf et al., 2005). With these limitations in mind, a degree of scepticism towards relating the results clinically is warranted, as some results may be driven by temporary physiological variations and food intake prior to blood testing. Further research is needed to investigate more novel aspects of cardiometabolic measures included in the study.

The longitudinal aspect of the study must also be discussed. Although the FreeSurfer longitudinal stream was carried out for both cross sectional and longitudinal data to avoid thickness estimates - and consequently T1 BAG - being influenced for follow up data, recent research suggests number of acquisitions per individual has an impact on the function of the FreeSurfer longitudinal pipeline (Beare et al., 2021). Moreover, the study may be limited due to a mean interval of 19.7 months for only one follow-up. Long-term longitudinal studies with several follow ups will be required to determine the temporal course and clinical predictive value of our findings in relation to future cardiometabolic disease and brain ageing.

The findings provide further support to the notion of the brain age gap reflecting individual variation in brain ageing (de Lange et al., 2020; Niu et al., 2019). While the modality-specific grey and white matter models showed similar performance, T1- and DTI-based BAGs revealed different associations with various CMRs. While it is likely that tissue specific brain age models capture biologically distinct information beyond single-modality models (de Lange et al., 2020; Richard et al., 2018; Smith et al., 2020), future research should look into additional regional modelling of tissue-specific brain ageing to detect associations with CMRs and other health indicators.

### 4.5. Conclusion

Our findings support that cardiometabolic risk factors including systolic blood pressure, WHR, and smoking, are associated with an older-appearing brain and accelerated brain ageing. While evidence demonstrating that effective management of modifiable CMRs reduces severity of associated brain imaging abnormalities is needed, promotion of improved cardiometabolic health and increasing existing knowledge on the links between the structure and function of the brain and cardiometabolic health can aid the development of preventative and risk-management treatment strategies in the general population and, likely, among patients with neurodegenerative and mental disorders.

## Data Availability

Data and code will be openly accessible on Open Science Framework.(https://osf.io/7z6cv/)

## 5. Acknowledgements

The study is supported by the Research Council of Norway (223273, 249795, 248238, 276082), the South-Eastern Norway Regional Health Authority (2014097, 2015044, 2015073, 2016083, 2018037, 2018076), the Norwegian ExtraFoundation for Health and Rehabilitation (2015/FO5146), KG Jebsen Stiftelsen, Swiss National Science Foundation (grant PZ00P3_193658), German Federal Ministry of Education and Research (BMBF, grant 01ZX1904A), ERA-Net Cofund through the ERA PerMed project ‘IMPLEMENT’ (Research Council of Norway – 298646), and the European Research Council under the European Union’s Horizon 2020 Research and Innovation program (ERC StG, Grant # 802998 and RIA Grant # 847776).

