## Supplementary Material for "Cardiometabolic risk factors associated with brain age and accelerate brain ageing"

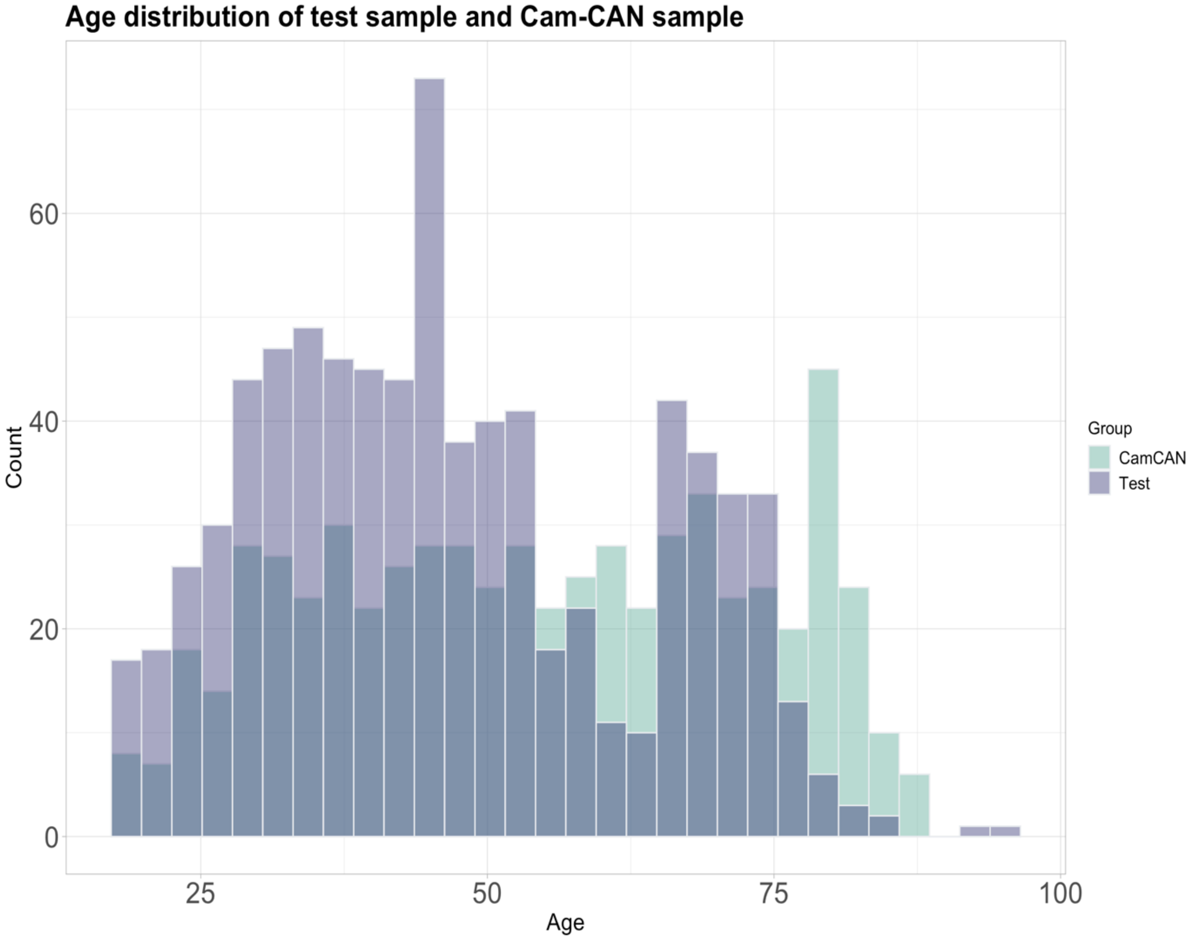


**SI Figure 1.** Age distribution for each sample. Cam-CAN cohort N = 622, StrokeTOP cohort N = 790.

**SI table 1.** Overview of Quality Assurance (QA) metrics for DTI data

| QA metric abbreviation | Measure |
|---|---|
| tsnr | Temporal-signal-to-noise-ratio |
| gmean | Global mean intensity |
| drift | Linear drift of signal over time |
| outmax | Outlier measurement maximum |
| outmean | Outlier measurement average |
| meanABSrmsb | Average absolute root-mean square |
| meanRELrmsb | Average relative root-mean square |
| maxABSrmsb | Maximum absolute root-mean square |
| maxRELrmsb | Maximum relative root-mean square |

**SI table 2.** Overview of Cardiometabolic risk factors (CMRs) used.

| Abbreviation used | Full description | Summary | Reference region (www.labhandbok.no) |
|---|---|---|---|
| BMI | Body mass index | Measure of body fat based on height and weight. | 18.5 – 24.9 |
| Smoking | Smoking | the action or habit of inhaling and exhaling the smoke of tobacco. | N/A |
| WHR | Waist-to-hip ratio | Dimensionless ratio of the circumference of the waist to that of the hips. | 0.81-0.85 (women), 0.96-1.0 (men) |
| Systolic | Systolic | The blood pressure when the heart is contracting. | 90-120 |
| Diastolic | Diastolic | The pressure in the arteries when the heart rests between beats. | 60-80 |
| Pulse | Pulse | The heart rate - number of times the heart beats in one minute. | 60-100 |
| Haemoglobin | Haemoglobin | The iron-containing oxygen-transport metalloprotein in the red blood cells. | 11,7 – 15,3 g/dL (women), 13,4 – 17,0 g/dL (men) |
| MVC | Mean corpuscular volume | Value that measures the average size and volume of a red blood cell. | 82 – 98 fL |
| MCH | Mean corpuscular haemoglobin | The average quantity of haemoglobin present in a single red blood cell. | 27 – 33 pg |
| MCHC | Mean corpuscular haemoglobin concentration | A measure of the average concentration of haemoglobin inside a single red blood cell. | 32 – 35 g/dL |
| Thrombocytes | Thrombocytes /Platelets | component of blood whose function is to react to bleeding by clumping, thereby initiating a blood clot. | 145 – 390 · 10^9/L |
| Sodium | Sodium | Sodium is an electrolyte that is key to helping send electrical signals between cells and controlling the amount of fluid in your body. | 137 – 145 mmol/L |
| Potassium | Potassium | High levels are dangerous and could be sign of hypercalcemia. Kidney failure is the most common cause of high potassium. | 3,6 -5,0 mmol/L |
| Chloride | Chloride | Chloride is a type of electrolyte. Electrolytes are electrically charged minerals that help control the amount of fluids and the balance of acids and bases in your body. | 97 – 110 mmol/L |
| Calcium | Calcium | If there is too much or too little calcium in the blood, it may be a sign of bone disease, thyroid disease, kidney disease, or other medical conditions. | 2,18 – 2,60 mmol/L |
| Magnesium | Magnesium | Magnesium deficiencies (hypomagnesemia) may be seen with malnutrition, conditions that cause malabsorption, and with excess loss of magnesium by the kidneys. Magnesium excess (hypermagnesemia) may be seen with the ingestion of antacids. | 0,71 – 1,08 mmol/L |
| Phosphate | Phosphate | Normally, the kidneys filter and remove excess phosphate from the blood. If phosphate levels in your blood are too high or too low, it can be a sign of kidney disease or other serious disorder. | 0,85 – 1,50 mmol/L (women), 0,75 – 1,65 mmol/L (men aged 18-49), 0,75 – 1,35 mmol/ (men aged 49+) |
| Creatinine | Creatinine | Normally, your kidneys filter creatinine from your blood and send it out of the body in your urine. If there is a problem with your | 45 – 90 µmol/L (women), 60 – 105 µmol/L (men) |

| | | | |
|---|---|---|---|
| | | kidneys, creatinine can build up in the blood and less will be released in urine. | |
| ALAT | Alanine transaminase | High levels of ALT in the blood can indicate a liver problem, even before you have signs of liver disease | ≤ 45 U/L (women), ≤ 70 U/L (men) |
| CK | Creatine kinase | CK is a type of protein, known as an enzyme. A small amount of CK in the blood is normal. Higher amounts can mean a health problem. | 35 – 210 U/L (women), 50 – 400 U/L (men aged 18-49), 40 – 280 U/L (men aged 49+) |
| LD | Lactate dehydrogenase | Higher than normal LD levels usually means you have some type of tissue damage or disease. Disorders that cause high LD levels include: Anemia, kidney disease, or liver disease. | 105– 205 U/L (aged 18-69), 115– 255 U/L (69+) |
| GT | Gamma-glutamyl transferase | GGT is elevated in the blood in most diseases that cause damage to the liver or bile ducts. | ≤ 45 U/L (women aged 18-39), ≤ 75 U/L (women aged 39+), ≤ 80 U/L (men aged 18-39), ≤ 115 U/L (men aged 39+) |
| CRP | C-reactive protein | CRP is a protein made by your liver. It's sent into your bloodstream in response to inflammation. High CRP is a marker of inflammation. | < 5 mg/L |
| Total cholesterol | Total cholesterol | a measure of the total amount of cholesterol in your blood. It includes both low-density lipoprotein (LDL) cholesterol and high-density lipoprotein (HDL) cholesterol. | Ages 18-29: 2,9 – 6,1 mmol/L, ages 30-49: 3,3 – 6,9 mmol/L, ages 49+: 3,9 – 7,8 mmol/L |
| HDL cholesterol | high-density lipoprotein cholesterol | known as the "good" cholesterol because it helps remove other forms of cholesterol from your bloodstream. Higher levels of HDL cholesterol are associated with a lower risk of heart disease. | 1,0 – 2,7 mmol/L (women), 0,8 – 2,1 mmol/L (men) |
| LDL cholesterol | low-density lipoprotein cholesterol | often called the "bad" cholesterol because it collects in the walls of your blood vessels, raising your chances of health problems like a heart attack or stroke. | Ages 18-29: 1,2 – 4,3 mmol/L, ages 30-49: 1,4 – 4,7 mmol/L, ages 49+: 2,0 – 5,3 mmol/L |
| Triglycerides | Triglycerides | type of fat (lipid) found in your blood. When you eat, your body converts any calories it doesn't need to use right away into triglycerides. | 0,45 – 2,60 mmol/L |
| Glucose | Glucose | Glucose is a type of sugar. A hormone called insulin helps move glucose from your bloodstream into your cells. High glucose may indicate diabetes. | 4,0 – 6,0 mmol/L |

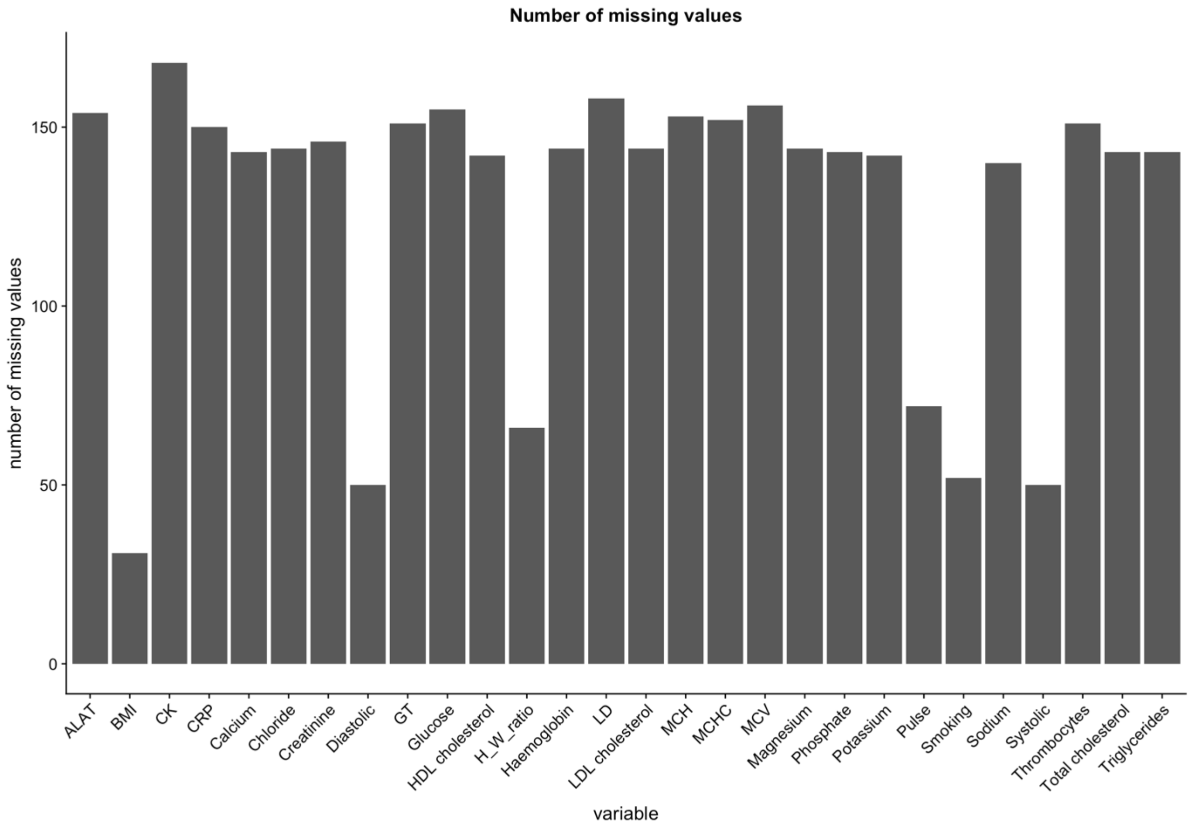


**SI Figure 2.** Showing number of missing values for each of the cardiometabolic risk factors used in the study.

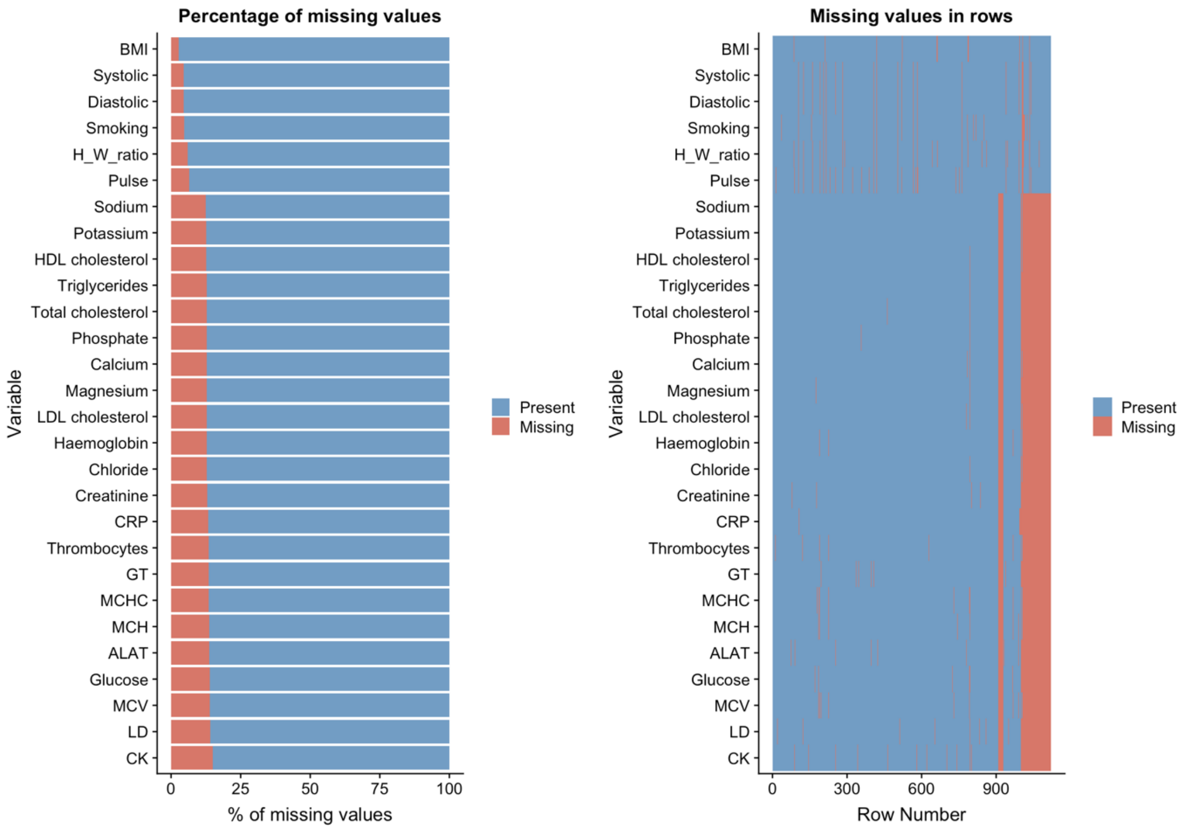


**SI Figure 3.** Showing percentage of missing data (left) and placement of missing data (right) for each cardiometabolic risk factor prior to imputation.

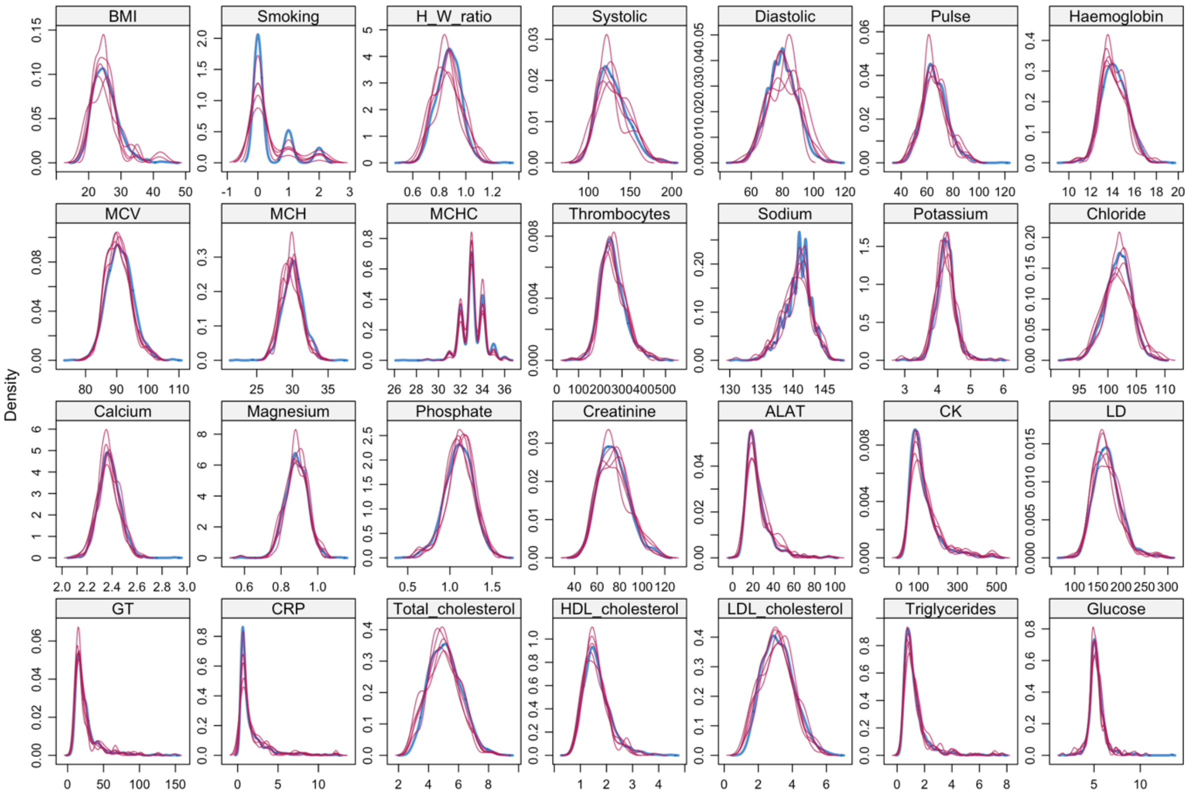


**SI Figure 4.** Showing density distribution of imputed cardiometabolic risk factors for five separate imputations. Blue lines represent original data, red lines represent imputed data.

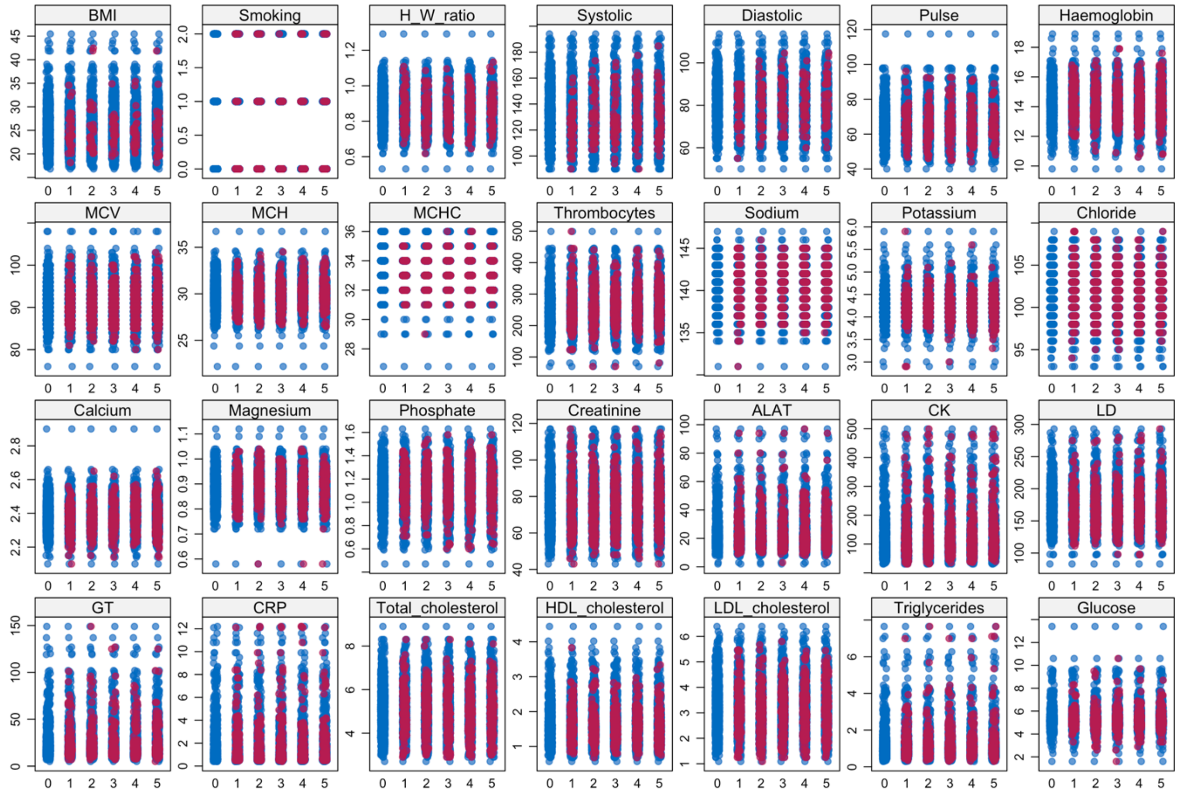


**SI Figure 5.** Showing strip plot of imputed cardiometabolic risk factors for five separate imputations. Blue dots represent original data, red dots represent imputed data.

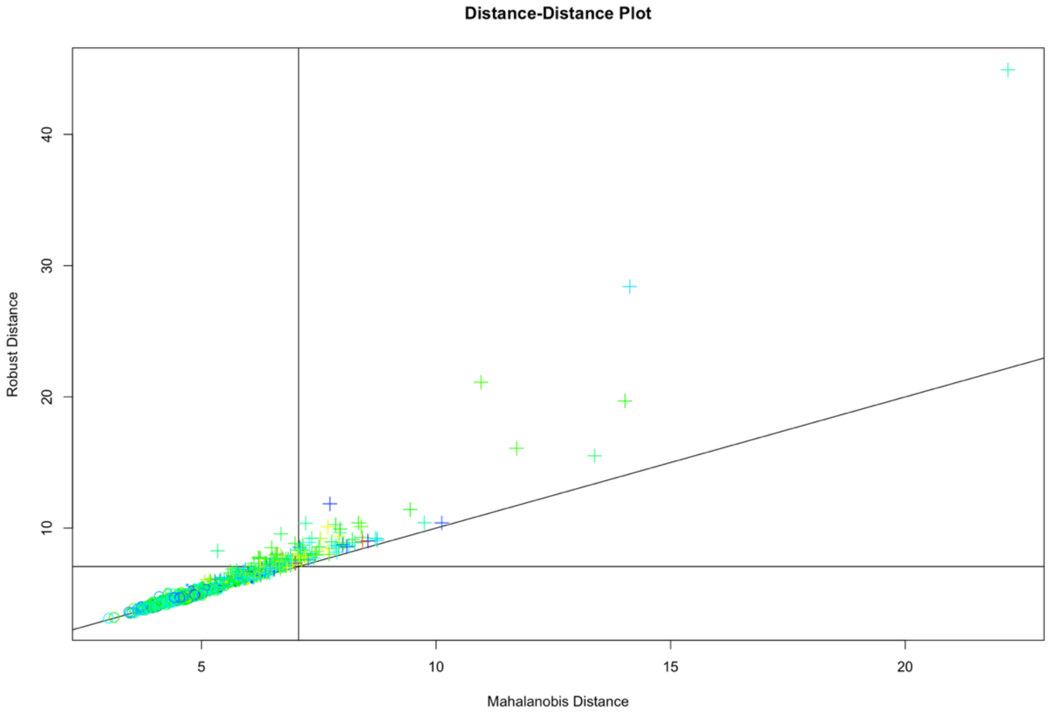


**SI Figure 6.** Showing the Mahalanobis distance plot, which measures the distance between a point and a distribution to which that point belongs.

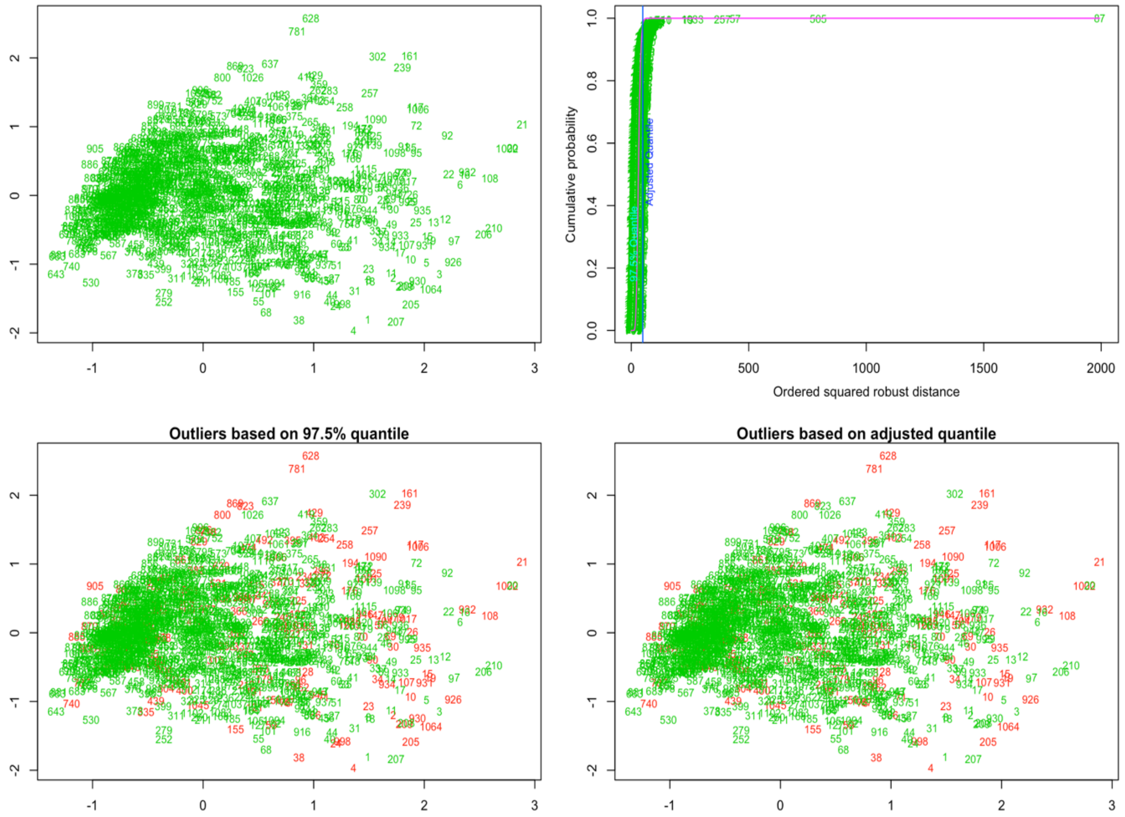


**SI Figure 7.** Showing the quantile and adjusted quantile plot, which will solve for and order the squared Mahalanobis Distances for the given observations and plot them against the empirical Chi-Squared distribution function of these values. Every observation outside of the Chi-Square quantile is coloured in red.

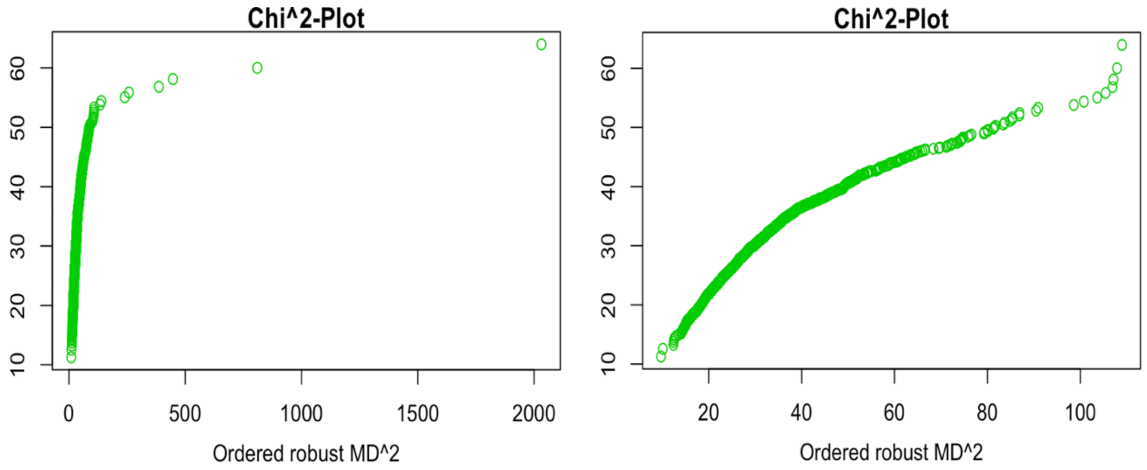


**SI Figure 8.** Showing the manual removal of outliers using an interactive plot, whereby the adjusted quantile threshold is set by the user, ideally to when the data resembles normal distribution. Here, eight outliers are removed from the left-hand side plot, resulting in the right-hand side plot.

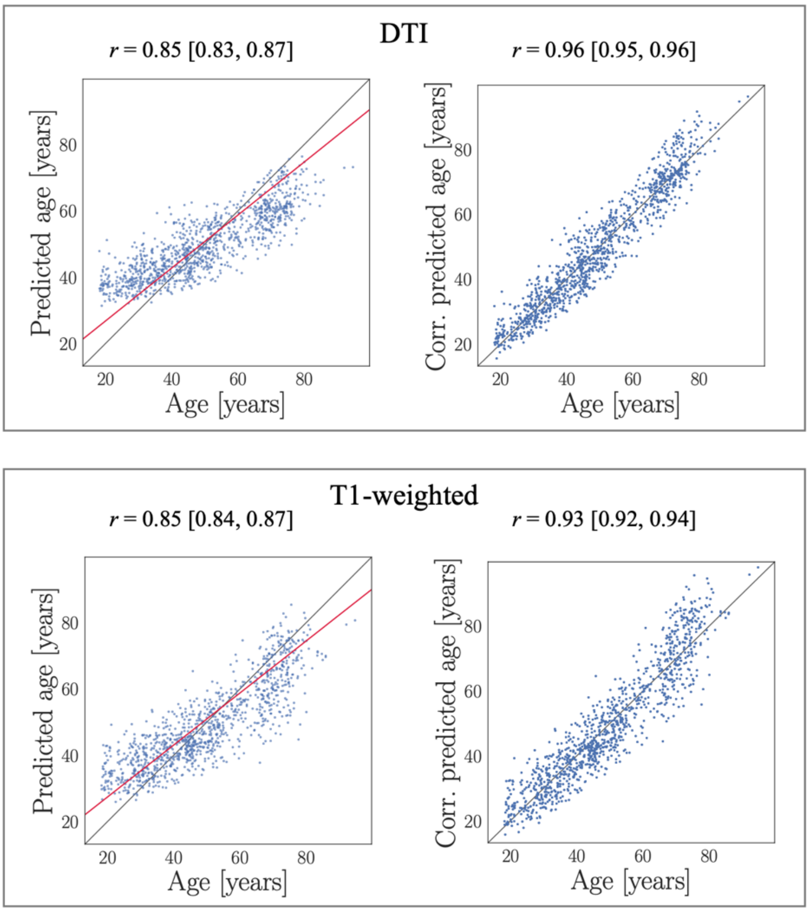


**SI Figure 9.** Age predictions before and after age-bias correction, where the plots on the left show the DTI and T1-weighed age predictions in the test sample. The red lines indicate the model fits in the Cam-CAN training sample, with overestimated predictions for younger participants and underestimated predictions for older participants. The plots on the right show the predictions after correcting for age-bias using the derived values of $\alpha$ and $\beta$ from the fit described in section 2.9 ($Y = \alpha \times \Omega + \beta$) with *Corrected Predicted Age* = *Predicted Age* + $[\Omega - (\alpha \times \Omega + \beta)]$.

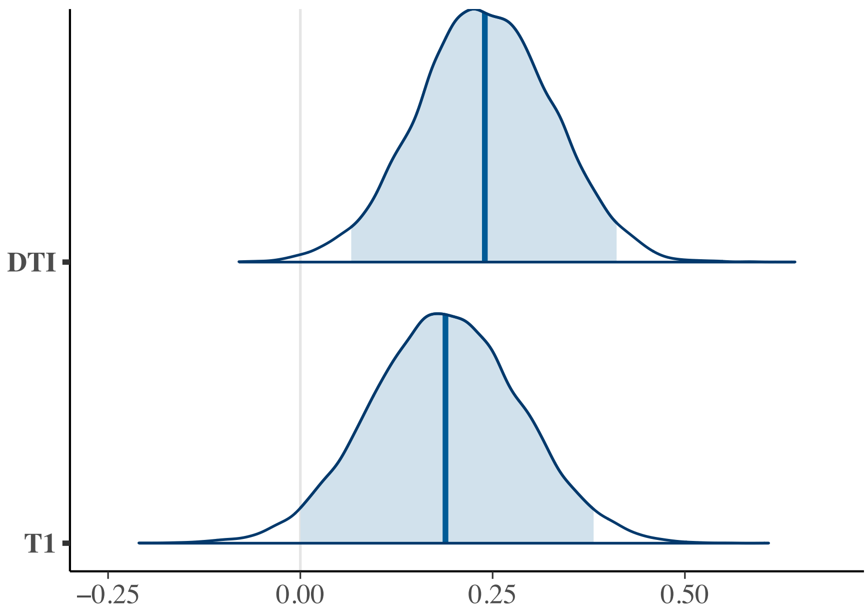


**SI Figure 10**. Posterior distributions of time on DTI and T1 BAGs. Shaded area represents 95% credible interval and vertical blue lines represent the mean of the posterior distribution.

**SI Table 3.** Showing Bayes factor (BF) interpretation.

| Bayes factor $BF_{12}$ | | | Interpretation |
|---|---|---|---|
| | > | 100 | Extreme evidence for $M_1$ |
| 30 | - | 100 | Very Strong evidence for $M_1$ |
| 10 | - | 30 | Strong evidence for $M_1$ |
| 3 | - | 10 | Moderate evidence for $M_1$ |
| 1 | - | 3 | Anecdotal evidence for $M_1$ |
| | 1 | | No evidence |
| 1/3 | - | 1 | Anecdotal evidence for $M_2$ |
| 1/10 | - | 1/3 | Moderate evidence for $M_2$ |
| 1/30 | - | 1/10 | Strong evidence for $M_2$ |
| 1/100 | - | 1/30 | Very Strong evidence for $M_2$ |
| | < | 1/100 | Extreme evidence for $M_2$ |

**SI Table 4.** Associations between cardiometabolic risk factors and time and age. CMR ~ Time + Sex + Age, and CMR ~ Age + Sex + Time (models = main effect).

| **CMR** | **predictor** | **Est.** | **lower95** | **upper95** | **p_higher** | **p_lower** | **evidence** |
|---|---|---|---|---|---|---|---|
| Pulse | TP | 0.008 | -0.041 | 0.055 | 0.624 | 0.376 | 11.78291 |
| Pulse | Age | 0.072 | 0.005 | 0.141 | 0.981 | 0.019 | 1.01927 |
| Thrombocytes | TP | -0.029 | -0.066 | 0.007 | 0.062 | 0.938 | 5.12962 |
| Thrombocytes | Age | -0.019 | -0.088 | 0.051 | 0.295 | 0.705 | 7.50634 |
| CRP | TP | 0.002 | -0.058 | 0.061 | 0.52 | 0.48 | 9.50071 |
| CRP | Age | 0.069 | 0.004 | 0.136 | 0.981 | 0.019 | 1.06579 |
| Phosphate | TP | 0.019 | -0.031 | 0.07 | 0.776 | 0.224 | 8.72517 |
| Phosphate | Age | -0.254 | -0.322 | -0.19 | 0 | 1 | 0 |

| | | | | | | | |
|---|---|---|---|---|---|---|---|
| Calcium | TP | -0.13 | -0.18 | -0.079 | 0 | 1 | 0 |
| Calcium | Age | 0.026 | -0.042 | 0.092 | 0.768 | 0.232 | 6.67769 |
| HDL_cholesterol | TP | -0.024 | -0.066 | 0.017 | 0.131 | 0.869 | 7.41746 |
| HDL_cholesterol | Age | 0.17 | 0.105 | 0.238 | 1 | 0 | 0 |
| Glucose | TP | -0.043 | -0.095 | 0.01 | 0.057 | 0.943 | 3.44237 |
| Glucose | Age | 0.251 | 0.186 | 0.315 | 1 | 0 | 0 |
| BMI | TP | 0.047 | 0.021 | 0.072 | 1 | 0 | 0.03025 |
| BMI | Age | 0.06 | -0.013 | 0.132 | 0.948 | 0.052 | 2.08499 |
| Triglycerides | TP | -0.078 | -0.127 | -0.032 | 0.001 | 0.999 | 0.08025 |
| Triglycerides | Age | 0.069 | -0.001 | 0.139 | 0.974 | 0.027 | 1.40695 |
| ALAT | TP | -0.034 | -0.089 | 0.023 | 0.119 | 0.881 | 5.3626 |
| ALAT | Age | 0.074 | 0.008 | 0.143 | 0.983 | 0.017 | 0.91255 |
| GT | TP | -0.048 | -0.092 | -0.002 | 0.018 | 0.982 | 1.46907 |
| GT | Age | 0.206 | 0.14 | 0.276 | 1 | 0 | 0 |
| WHR | TP | -0.104 | -0.145 | -0.063 | 0 | 1 | 0 |
| WHR | Age | 0.448 | 0.385 | 0.513 | 1 | 0 | 0 |
| Haemoglobin | TP | -0.067 | -0.107 | -0.026 | 0.001 | 0.999 | 0.06782 |
| Haemoglobin | Age | 0.091 | 0.022 | 0.161 | 0.993 | 0.007 | 0.35027 |
| CK | TP | -0.014 | -0.066 | 0.034 | 0.296 | 0.704 | 9.82579 |
| CK | Age | -0.065 | -0.134 | 0.005 | 0.032 | 0.968 | 1.615 |
| LD | TP | 0.058 | 0.012 | 0.107 | 0.993 | 0.007 | 0.63163 |
| LD | Age | 0.328 | 0.262 | 0.39 | 1 | 0 | 0 |
| MCHC | TP | -0.019 | -0.074 | 0.034 | 0.24 | 0.76 | 8.59256 |
| MCHC | Age | -0.106 | -0.17 | -0.037 | 0.001 | 0.999 | 0.07687 |
| Creatinine | TP | -0.05 | -0.093 | -0.006 | 0.011 | 0.989 | 1.1259 |
| Creatinine | Age | 0.049 | -0.021 | 0.117 | 0.916 | 0.084 | 3.24273 |
| Magnesium | TP | 0.003 | -0.052 | 0.056 | 0.543 | 0.457 | 10.75853 |
| Magnesium | Age | 0.067 | -0.002 | 0.133 | 0.974 | 0.026 | 1.28759 |
| Sodium | TP | 0.135 | 0.076 | 0.192 | 1 | 0 | 0.00194 |
| Sodium | Age | 0.114 | 0.051 | 0.18 | 1 | 0 | 0.03449 |
| Chloride | TP | 0.149 | 0.093 | 0.207 | 1 | 0 | 0 |
| Chloride | Age | -0.031 | -0.096 | 0.034 | 0.176 | 0.824 | 5.81501 |
| MCV | TP | 0.095 | 0.052 | 0.138 | 1 | 0 | 0.0007 |
| MCV | Age | 0.298 | 0.236 | 0.364 | 1 | 0 | 0 |
| MCH | TP | 0.074 | 0.028 | 0.119 | 0.999 | 0.001 | 0.08411 |
| MCH | Age | 0.213 | 0.148 | 0.281 | 1 | 0 | 0 |
| Total_cholesterol | TP | -0.072 | -0.121 | -0.019 | 0.003 | 0.997 | 0.28113 |
| Total_cholesterol | Age | 0.358 | 0.295 | 0.421 | 1 | 0 | 0 |
| LDL_cholesterol | TP | -0.007 | -0.061 | 0.043 | 0.394 | 0.606 | 10.70187 |
| LDL_cholesterol | Age | 0.301 | 0.236 | 0.364 | 1 | 0 | 0 |
| Systolic | TP | -0.077 | -0.114 | -0.044 | 0 | 1 | 0.00002 |
| Systolic | Age | 0.589 | 0.533 | 0.644 | 1 | 0 | 0 |
| Diastolic | TP | -0.134 | -0.174 | -0.096 | 0 | 1 | 0 |
| Diastolic | Age | 0.329 | 0.26 | 0.394 | 1 | 0 | 0 |
| Potassium | TP | -0.06 | -0.114 | -0.004 | 0.017 | 0.983 | 1.02352 |
| Potassium | Age | 0.179 | 0.112 | 0.246 | 1 | 0 | 0 |
| Smoking | TP | -0.055 | -0.094 | -0.016 | 0.004 | 0.996 | 0.37051 |
| Smoking | Age | 0.229 | 0.16 | 0.296 | 1 | 0 | 0 |

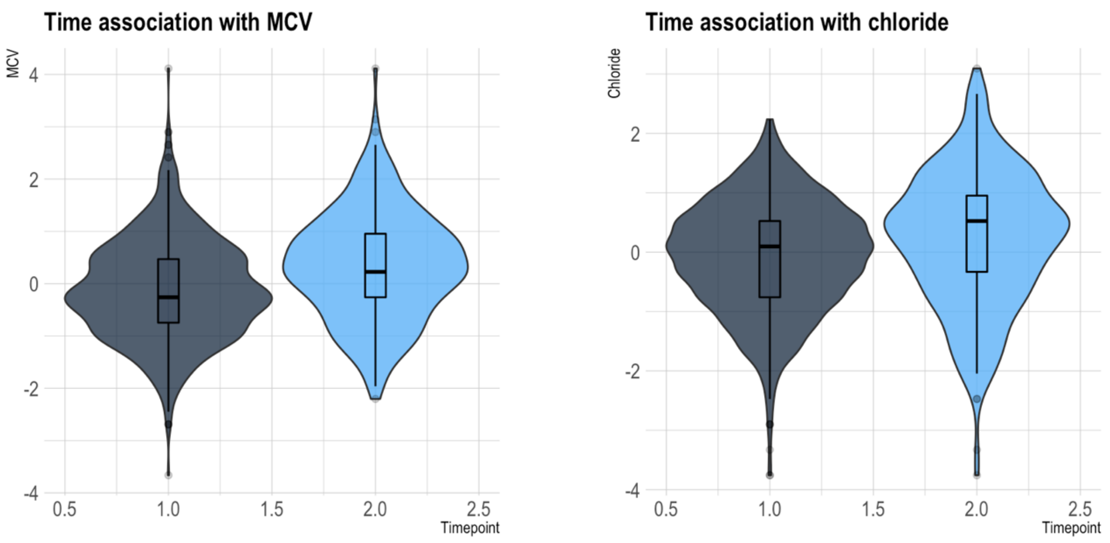


**SI Figure 11. Associations between time and a selection of CMH indicators.** Violin plots with box plots demonstrating the increase in CMH indicator measurement at follow up (light blue) compared to baseline. (dark blue).

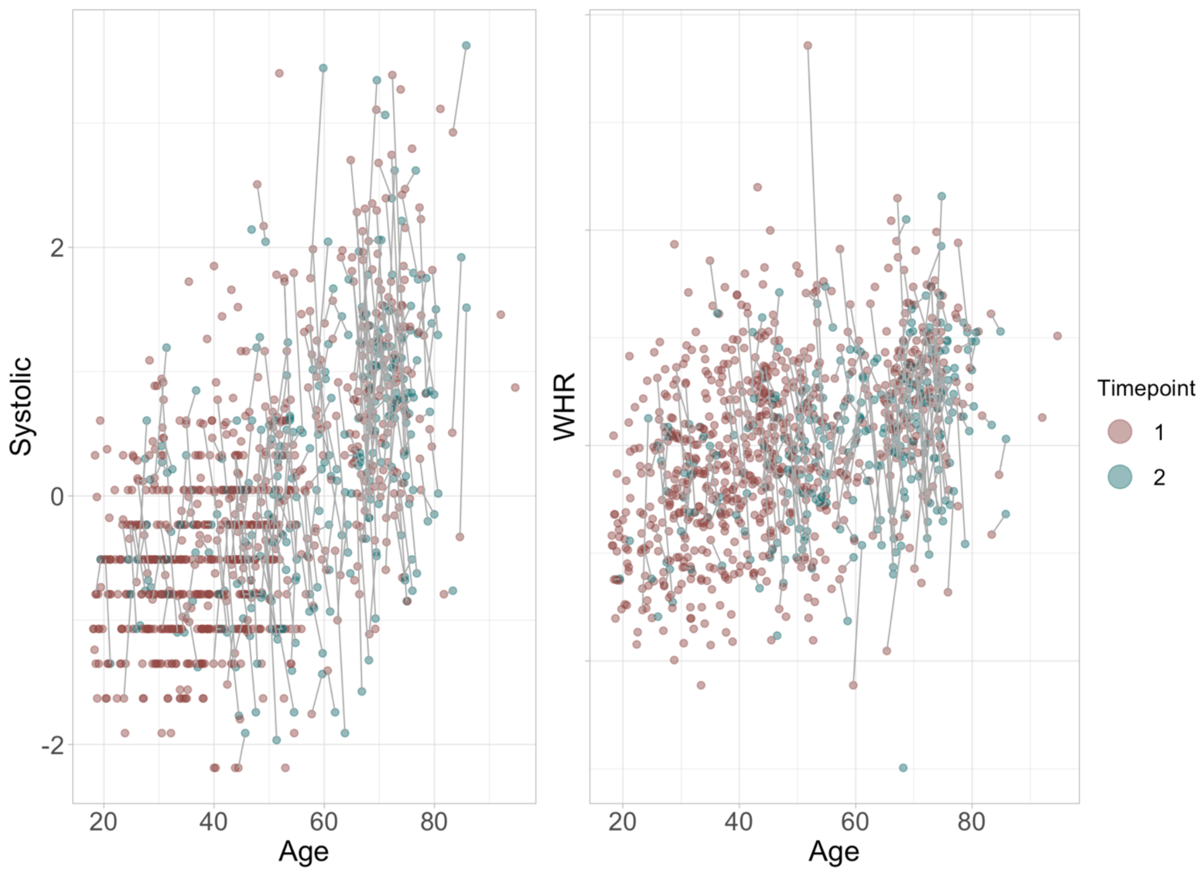


**SI Figure 12. Systolic blood pressure and WHR as functions of age.** The figure shows baseline measures in red and follow up measures in green.

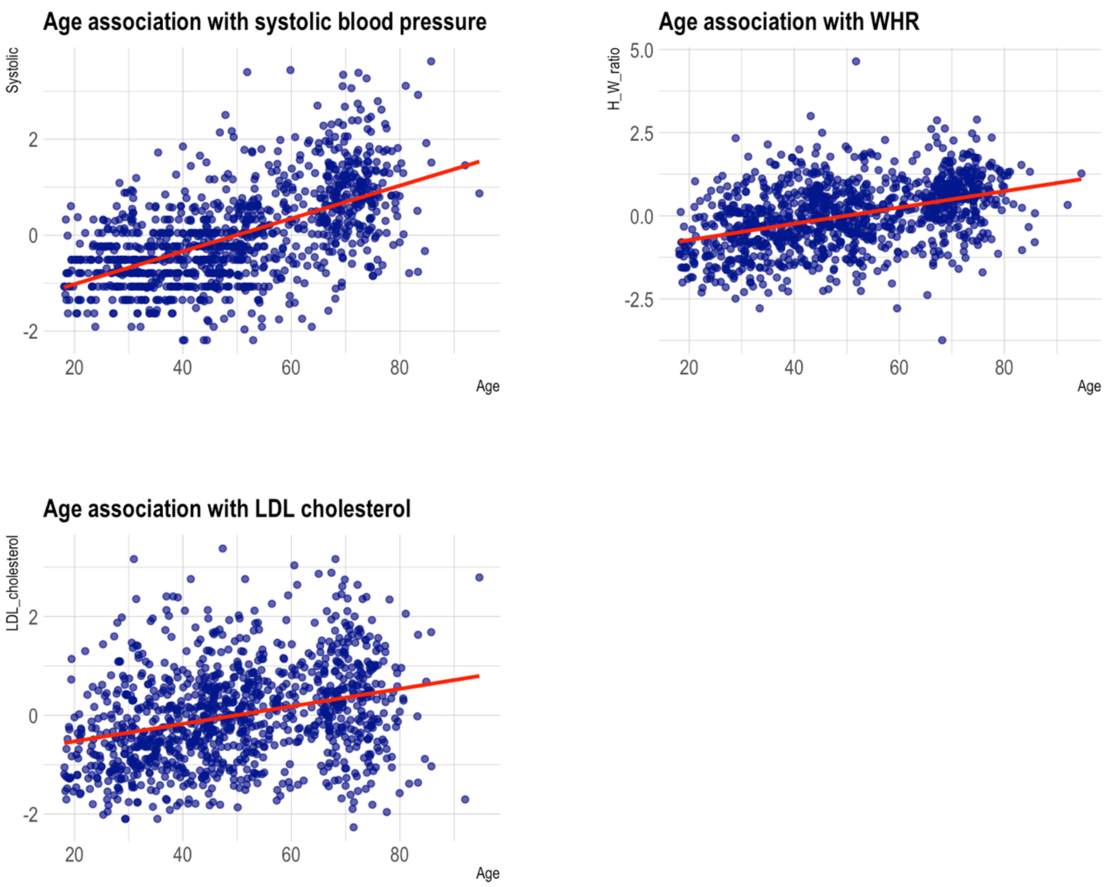


**SI Figure 13. Associations between age and a selection of CMH indicators.** Scatter plots demonstrating the increase in CMH indicator measurement with increasing age.

**SI Table 5.** DTI BAG ~ CMR + Sex + TP (model = main effect)

| CMR | Est. | lower95 | upper95 | p_higher | p_lower | evidence | prior |
|---|---|---|---|---|---|---|---|
| Pulse | -0.03 | -0.28 | 0.22 | 0.41 | 0.59 | 2.35 | 0.3 |
| Thrombocytes | 0 | -0.27 | 0.27 | 0.51 | 0.49 | 2.18 | 0.3 |
| CRP | 0.03 | -0.2 | 0.25 | 0.59 | 0.41 | 2.53 | 0.3 |
| Phosphate | 0.29 | 0.04 | 0.52 | 0.99 | 0.01 | 0.17 | 0.3 |
| Calcium | 0.14 | -0.1 | 0.39 | 0.86 | 0.14 | 1.39 | 0.3 |
| HDL_cholesterol | -0.16 | -0.42 | 0.11 | 0.11 | 0.89 | 1.11 | 0.3 |
| Glucose | -0.14 | -0.38 | 0.1 | 0.12 | 0.88 | 1.25 | 0.3 |
| BMI | -0.13 | -0.4 | 0.16 | 0.19 | 0.81 | 1.4 | 0.3 |
| Triglycerides | -0.03 | -0.28 | 0.21 | 0.41 | 0.59 | 2.31 | 0.3 |
| ALAT | -0.05 | -0.29 | 0.19 | 0.34 | 0.66 | 2.37 | 0.3 |
| GT | 0.1 | -0.15 | 0.36 | 0.78 | 0.22 | 1.78 | 0.3 |
| WHR | 0.17 | -0.09 | 0.43 | 0.9 | 0.1 | 0.89 | 0.3 |
| Haemoglobin | -0.04 | -0.3 | 0.23 | 0.38 | 0.62 | 2.14 | 0.3 |
| CK | -0.06 | -0.31 | 0.17 | 0.3 | 0.7 | 2.17 | 0.3 |
| LD | -0.14 | -0.37 | 0.13 | 0.15 | 0.85 | 1.38 | 0.3 |
| MCHC | 0.05 | -0.18 | 0.29 | 0.66 | 0.34 | 2.3 | 0.3 |
| Creatinine | 0.23 | -0.03 | 0.48 | 0.96 | 0.04 | 0.6 | 0.3 |
| Magnesium | -0.11 | -0.34 | 0.13 | 0.19 | 0.81 | 1.62 | 0.3 |
| Sodium | 0.09 | -0.14 | 0.32 | 0.79 | 0.21 | 1.79 | 0.3 |

| | | | | | | | |
|---|---|---|---|---|---|---|---|
| Chloride | -0.1 | -0.33 | 0.12 | 0.2 | 0.8 | 1.82 | 0.3 |
| MCV | -0.32 | -0.58 | -0.06 | 0.01 | 0.99 | 0.14 | 0.3 |
| MCH | -0.24 | -0.5 | 0.02 | 0.03 | 0.97 | 0.46 | 0.3 |
| Total_cholesterol | -0.19 | -0.42 | 0.06 | 0.07 | 0.93 | 0.76 | 0.3 |
| LDL_cholesterol | -0.1 | -0.34 | 0.13 | 0.2 | 0.8 | 1.72 | 0.3 |
| Systolic | 0.15 | -0.14 | 0.41 | 0.85 | 0.15 | 1.26 | 0.3 |
| Diastolic | 0.02 | -0.23 | 0.3 | 0.56 | 0.44 | 2.21 | 0.3 |
| Potassium | -0.01 | -0.25 | 0.22 | 0.45 | 0.55 | 2.55 | 0.3 |
| Smoking | -0.03 | -0.3 | 0.23 | 0.41 | 0.59 | 2.14 | 0.3 |

**SI Table 6.** T1 BAG ~ CMR + Sex + TP (model = main effect)

| CMR | Est. | lower95 | upper95 | p_higher | p_lower | evidence | prior |
|---|---|---|---|---|---|---|---|
| Pulse | 0.29 | -0.01 | 0.58 | 0.97 | 0.03 | 0.3 | 0.3 |
| Thrombocytes | 0.13 | -0.18 | 0.45 | 0.79 | 0.21 | 1.35 | 0.3 |
| CRP | 0.29 | 0.03 | 0.54 | 0.99 | 0.01 | 0.21 | 0.3 |
| Phosphate | -0.04 | -0.32 | 0.24 | 0.4 | 0.6 | 1.95 | 0.3 |
| Calcium | 0.03 | -0.26 | 0.31 | 0.58 | 0.42 | 2.05 | 0.3 |
| HDL_cholesterol | 0.01 | -0.29 | 0.32 | 0.53 | 0.47 | 1.93 | 0.3 |
| Glucose | 0.08 | -0.19 | 0.35 | 0.72 | 0.28 | 1.88 | 0.3 |
| BMI | -0.2 | -0.53 | 0.12 | 0.11 | 0.89 | 0.86 | 0.3 |
| Triglycerides | -0.01 | -0.31 | 0.28 | 0.48 | 0.52 | 1.99 | 0.3 |
| ALAT | -0.05 | -0.33 | 0.23 | 0.37 | 0.63 | 1.93 | 0.3 |
| GT | 0.06 | -0.23 | 0.36 | 0.65 | 0.35 | 1.79 | 0.3 |
| WHR | -0.12 | -0.43 | 0.18 | 0.22 | 0.78 | 1.4 | 0.3 |
| Haemoglobin | -0.04 | -0.34 | 0.27 | 0.39 | 0.61 | 1.86 | 0.3 |
| CK | -0.08 | -0.37 | 0.19 | 0.3 | 0.7 | 1.73 | 0.3 |
| LD | 0.24 | -0.07 | 0.51 | 0.95 | 0.05 | 0.55 | 0.3 |
| MCHC | -0.11 | -0.38 | 0.16 | 0.21 | 0.79 | 1.63 | 0.3 |
| Creatinine | -0.26 | -0.55 | 0.05 | 0.05 | 0.95 | 0.52 | 0.3 |
| Magnesium | 0.02 | -0.24 | 0.3 | 0.57 | 0.43 | 2.15 | 0.3 |
| Sodium | 0 | -0.27 | 0.25 | 0.49 | 0.51 | 2.22 | 0.3 |
| Chloride | -0.17 | -0.44 | 0.09 | 0.1 | 0.9 | 1.03 | 0.3 |
| MCV | 0.02 | -0.29 | 0.31 | 0.54 | 0.46 | 1.92 | 0.3 |
| MCH | -0.03 | -0.32 | 0.27 | 0.43 | 0.57 | 1.94 | 0.3 |
| Total_cholesterol | -0.16 | -0.43 | 0.11 | 0.13 | 0.87 | 1.14 | 0.3 |
| LDL_cholesterol | -0.16 | -0.43 | 0.11 | 0.13 | 0.87 | 1.13 | 0.3 |
| Systolic | 0.37 | 0.06 | 0.69 | 0.99 | 0.01 | 0.13 | 0.3 |
| Diastolic | 0.03 | -0.27 | 0.34 | 0.59 | 0.41 | 1.85 | 0.3 |
| Potassium | 0.13 | -0.14 | 0.41 | 0.82 | 0.18 | 1.36 | 0.3 |
| Smoking | 0.35 | 0.04 | 0.66 | 0.99 | 0.01 | 0.17 | 0.3 |

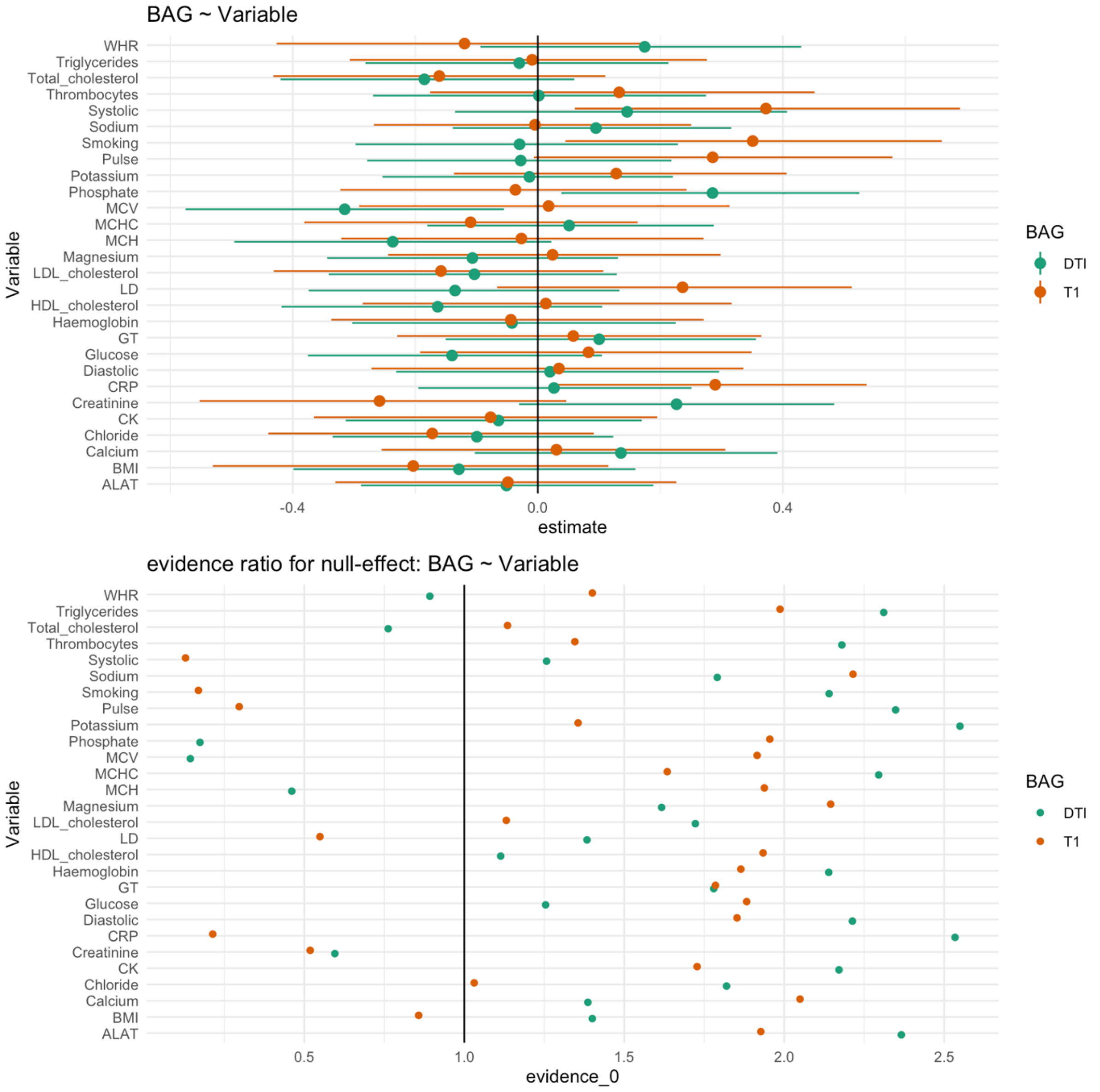


**SI Figure 14**. Estimates with 95% credible interval are shown in the upper figure and evidence ratios are shown in the lower figure. Values above 1 indicate evidence in favour of the effect being null and values below 1 indicate evidence in favour of an effect.

**SI Table 7.** DTI BAG ~ Time*CMR + Sex (model = interaction effect of time)

| Time:CMR | Est. | lower95 | upper95 | p_higher | p_lower | evidence | prior |
|---|---|---|---|---|---|---|---|
| TP:Pulse | 0.1 | -0.08 | 0.26 | 0.87 | 0.13 | 1.76 | 0.3 |
| TP:Thrombocytes | -0.02 | -0.18 | 0.15 | 0.4 | 0.6 | 3.34 | 0.3 |
| TP:CRP | -0.01 | -0.2 | 0.17 | 0.45 | 0.55 | 3.18 | 0.3 |
| TP:Phosphate | -0.03 | -0.21 | 0.15 | 0.38 | 0.62 | 3.11 | 0.3 |
| TP:Calcium | -0.09 | -0.28 | 0.1 | 0.17 | 0.83 | 1.97 | 0.3 |
| TP:HDL_cholesterol | -0.06 | -0.24 | 0.1 | 0.23 | 0.77 | 2.58 | 0.3 |
| TP:Glucose | -0.04 | -0.22 | 0.13 | 0.33 | 0.67 | 2.96 | 0.3 |
| TP:BMI | 0.06 | -0.11 | 0.23 | 0.76 | 0.24 | 2.63 | 0.3 |
| TP:Triglycerides | 0.09 | -0.14 | 0.3 | 0.78 | 0.22 | 1.98 | 0.3 |
| TP:ALAT | -0.02 | -0.22 | 0.18 | 0.44 | 0.56 | 3.06 | 0.3 |
| TP:GT | 0.05 | -0.14 | 0.23 | 0.69 | 0.31 | 2.82 | 0.3 |
| TP:WHR | 0.25 | 0.06 | 0.44 | 1 | 0 | 0.09 | 0.3 |

| | | | | | | | |
|---|---|---|---|---|---|---|---|
| TP:Haemoglobin | 0.03 | -0.15 | 0.21 | 0.64 | 0.36 | 2.94 | 0.3 |
| TP:CK | -0.01 | -0.19 | 0.17 | 0.45 | 0.55 | 3.23 | 0.3 |
| TP:LD | -0.01 | -0.18 | 0.18 | 0.46 | 0.54 | 3.2 | 0.3 |
| TP:MCHC | -0.05 | -0.22 | 0.14 | 0.31 | 0.69 | 2.86 | 0.3 |
| TP:Creatinine | 0.15 | -0.01 | 0.32 | 0.96 | 0.04 | 0.73 | 0.3 |
| TP:Magnesium | 0.06 | -0.11 | 0.25 | 0.75 | 0.25 | 2.58 | 0.3 |
| TP:Sodium | 0 | -0.19 | 0.19 | 0.51 | 0.49 | 3.1 | 0.3 |
| TP:Chloride | 0.04 | -0.15 | 0.21 | 0.66 | 0.34 | 3.16 | 0.3 |
| TP:MCV | -0.05 | -0.21 | 0.12 | 0.3 | 0.7 | 2.99 | 0.3 |
| TP:MCH | -0.07 | -0.23 | 0.11 | 0.2 | 0.8 | 2.46 | 0.3 |
| TP:Total_cholesterol | -0.03 | -0.21 | 0.15 | 0.36 | 0.64 | 3.14 | 0.3 |
| TP:LDL_cholesterol | -0.01 | -0.2 | 0.17 | 0.45 | 0.55 | 3.19 | 0.3 |
| TP:Systolic | 0.25 | 0.08 | 0.41 | 1 | 0 | 0.07 | 0.3 |
| TP:Diastolic | 0.1 | -0.07 | 0.28 | 0.88 | 0.12 | 1.71 | 0.3 |
| TP:Potassium | 0.08 | -0.1 | 0.27 | 0.81 | 0.19 | 2.25 | 0.3 |
| TP:Smoking | 0.19 | 0.02 | 0.37 | 0.99 | 0.01 | 0.31 | 0.3 |

**SI Table 8.** T1 BAG ~ Time*CMR + Sex (model = interaction effect of time)

| **Time:CMR** | **Est.** | **lower95** | **upper95** | **p_higher** | **p_lower** | **evidence** | **prior** |
|---|---|---|---|---|---|---|---|
| TP:Pulse | 0.17 | -0.01 | 0.37 | 0.96 | 0.04 | 0.63 | 0.3 |
| TP:Thrombocytes | -0.04 | -0.23 | 0.15 | 0.32 | 0.68 | 2.84 | 0.3 |
| TP:CRP | 0 | -0.2 | 0.21 | 0.5 | 0.5 | 2.73 | 0.3 |
| TP:Phosphate | -0.07 | -0.28 | 0.13 | 0.25 | 0.75 | 2.19 | 0.3 |
| TP:Calcium | -0.09 | -0.31 | 0.11 | 0.21 | 0.79 | 2 | 0.3 |
| TP:HDL_cholesterol | -0.11 | -0.31 | 0.07 | 0.12 | 0.88 | 1.52 | 0.3 |
| TP:Glucose | 0.04 | -0.16 | 0.24 | 0.66 | 0.34 | 2.64 | 0.3 |
| TP:BMI | 0.04 | -0.15 | 0.24 | 0.67 | 0.33 | 2.69 | 0.3 |
| TP:Triglycerides | 0.22 | -0.04 | 0.47 | 0.95 | 0.05 | 0.58 | 0.3 |
| TP:ALAT | 0.23 | 0.01 | 0.45 | 0.98 | 0.02 | 0.37 | 0.3 |
| TP:GT | 0.27 | 0.06 | 0.47 | 1 | 0 | 0.11 | 0.3 |
| TP:WHR | 0.3 | 0.09 | 0.51 | 1 | 0 | 0.06 | 0.3 |
| TP:Haemoglobin | 0.18 | -0.02 | 0.38 | 0.96 | 0.04 | 0.64 | 0.3 |
| TP:CK | -0.01 | -0.22 | 0.19 | 0.46 | 0.54 | 2.94 | 0.3 |
| TP:LD | 0.11 | -0.08 | 0.31 | 0.87 | 0.13 | 1.61 | 0.3 |
| TP:MCHC | -0.01 | -0.21 | 0.19 | 0.47 | 0.53 | 2.77 | 0.3 |
| TP:Creatinine | -0.07 | -0.26 | 0.13 | 0.24 | 0.76 | 2.32 | 0.3 |
| TP:Magnesium | 0.07 | -0.13 | 0.28 | 0.75 | 0.25 | 2.25 | 0.3 |
| TP:Sodium | -0.03 | -0.24 | 0.19 | 0.39 | 0.61 | 2.58 | 0.3 |
| TP:Chloride | -0.07 | -0.27 | 0.13 | 0.24 | 0.76 | 2.32 | 0.3 |
| TP:MCV | 0 | -0.2 | 0.19 | 0.49 | 0.51 | 3.01 | 0.3 |
| TP:MCH | -0.02 | -0.22 | 0.16 | 0.4 | 0.6 | 2.88 | 0.3 |
| TP:Total_cholesterol | 0.17 | -0.02 | 0.38 | 0.95 | 0.05 | 0.72 | 0.3 |
| TP:LDL_cholesterol | 0.21 | 0.01 | 0.42 | 0.98 | 0.02 | 0.38 | 0.3 |
| TP:Systolic | 0.05 | -0.14 | 0.25 | 0.7 | 0.3 | 2.54 | 0.3 |
| TP:Diastolic | 0.07 | -0.13 | 0.27 | 0.75 | 0.25 | 2.29 | 0.3 |
| TP:Potassium | -0.06 | -0.28 | 0.14 | 0.28 | 0.72 | 2.43 | 0.3 |
| TP:Smoking | 0.04 | -0.15 | 0.23 | 0.66 | 0.34 | 2.89 | 0.3 |

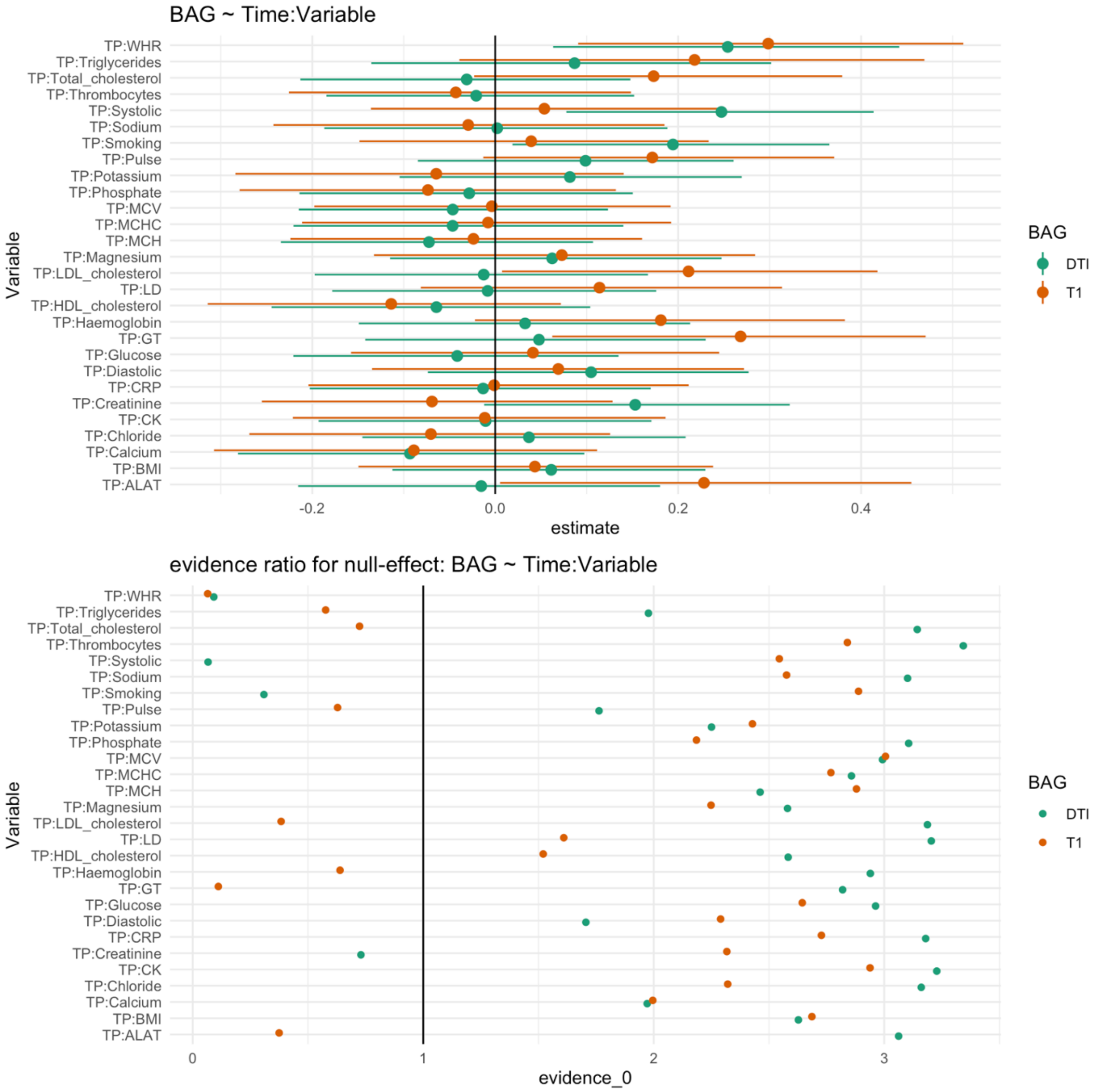


**SI Figure 15.** Estimates with 95% credible interval are shown in the upper figure and evidence ratios are shown in the lower figure. Values above 1 indicate evidence in favour of the effect being null and values below 1 indicate evidence in favour of an effect.

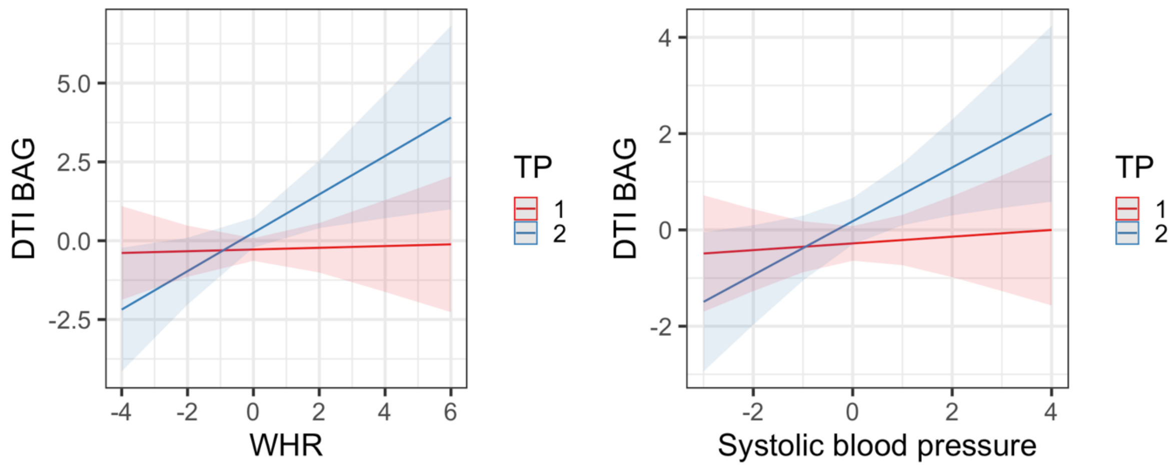


**SI Figure 16.** Interaction effects of CMH indicator variables and time on DTI BAG, where increases in BAG are seen with increases in WHR and systolic blood pressure over time.

**SI Table 9.** DTI BAG ~ Age*CMR + Sex (model = interaction effect of age)

| Age:CMR | Est. | lower95 | upper95 | p_higher | p_lower | evidence | prior |
|---|---|---|---|---|---|---|---|
| Age:Pulse | 0.13 | -0.12 | 0.37 | 0.85 | 0.15 | 1.48 | 0.3 |
| Age:Thrombocytes | -0.23 | -0.5 | 0.04 | 0.05 | 0.95 | 0.56 | 0.3 |
| Age:CRP | 0.05 | -0.18 | 0.25 | 0.68 | 0.32 | 2.51 | 0.3 |
| Age:Phosphate | -0.13 | -0.38 | 0.13 | 0.15 | 0.85 | 1.48 | 0.3 |
| Age:Calcium | 0.1 | -0.14 | 0.33 | 0.8 | 0.2 | 1.69 | 0.3 |
| Age:HDL_cholesterol | 0.11 | -0.19 | 0.39 | 0.77 | 0.23 | 1.54 | 0.3 |
| Age:Glucose | 0.24 | 0.01 | 0.48 | 0.98 | 0.02 | 0.36 | 0.3 |
| Age:BMI | 0.21 | -0.08 | 0.49 | 0.93 | 0.07 | 0.72 | 0.3 |
| Age:Triglycerides | 0.02 | -0.27 | 0.32 | 0.56 | 0.44 | 1.99 | 0.3 |
| Age:ALAT | 0.14 | -0.11 | 0.38 | 0.86 | 0.14 | 1.28 | 0.3 |
| Age:GT | 0.36 | 0.09 | 0.63 | 0.99 | 0.01 | 0.09 | 0.3 |
| Age:WHR | 0.31 | 0.04 | 0.57 | 0.99 | 0.01 | 0.18 | 0.3 |
| Age:Haemoglobin | 0.07 | -0.2 | 0.33 | 0.68 | 0.32 | 1.98 | 0.3 |
| Age:CK | -0.25 | -0.52 | 0 | 0.03 | 0.97 | 0.38 | 0.3 |
| Age:LD | 0.04 | -0.2 | 0.28 | 0.63 | 0.37 | 2.29 | 0.3 |
| Age:MCHC | -0.07 | -0.33 | 0.17 | 0.29 | 0.71 | 2.08 | 0.3 |
| Age:Creatinine | -0.25 | -0.5 | 0.01 | 0.03 | 0.97 | 0.4 | 0.3 |
| Age:Magnesium | 0 | -0.24 | 0.24 | 0.49 | 0.51 | 2.39 | 0.3 |
| Age:Sodium | 0.02 | -0.2 | 0.27 | 0.58 | 0.42 | 2.46 | 0.3 |
| Age:Chloride | -0.01 | -0.24 | 0.23 | 0.46 | 0.54 | 2.52 | 0.3 |
| Age:MCV | 0.1 | -0.16 | 0.35 | 0.78 | 0.22 | 1.77 | 0.3 |
| Age:MCH | 0.04 | -0.21 | 0.3 | 0.63 | 0.37 | 2.25 | 0.3 |
| Age:Total_cholesterol | 0.14 | -0.09 | 0.39 | 0.88 | 0.12 | 1.25 | 0.3 |
| Age:LDL_cholesterol | 0.15 | -0.1 | 0.38 | 0.89 | 0.11 | 1.22 | 0.3 |
| Age:Systolic | 0.44 | 0.15 | 0.72 | 1 | 0 | 0.02 | 0.3 |
| Age:Diastolic | 0.23 | -0.03 | 0.51 | 0.95 | 0.05 | 0.51 | 0.3 |
| Age:Potassium | -0.2 | -0.43 | 0.04 | 0.05 | 0.95 | 0.6 | 0.3 |
| Age:Smoking | 0.04 | -0.24 | 0.31 | 0.6 | 0.4 | 2.1 | 0.3 |

**SI Table 10.** T1 BAG ~ Age*CMR + Sex (model = interaction effect of age)

| **Age:CMR** | **estimate** | **lower95** | **upper95** | **p_higher** | **p_lower** | **evidence** | **prior** |
|---|---|---|---|---|---|---|---|
| Age:Pulse | 0.3 | 0.02 | 0.57 | 0.98 | 0.02 | 0.25 | 0.3 |
| Age:Thrombocytes | -0.06 | -0.38 | 0.25 | 0.35 | 0.65 | 1.74 | 0.3 |
| Age:CRP | 0.42 | 0.18 | 0.66 | 1 | 0 | 0.01 | 0.3 |
| Age:Phosphate | -0.23 | -0.51 | 0.07 | 0.06 | 0.94 | 0.63 | 0.3 |
| Age:Calcium | 0.1 | -0.17 | 0.37 | 0.76 | 0.24 | 1.61 | 0.3 |
| Age:HDL_cholesterol | -0.15 | -0.47 | 0.19 | 0.2 | 0.8 | 1.2 | 0.3 |
| Age:Glucose | 0.33 | 0.06 | 0.61 | 0.99 | 0.01 | 0.15 | 0.3 |
| Age:BMI | 0.24 | -0.09 | 0.58 | 0.92 | 0.08 | 0.67 | 0.3 |
| Age:Triglycerides | 0.36 | 0.02 | 0.69 | 0.98 | 0.02 | 0.19 | 0.3 |
| Age:ALAT | 0.25 | -0.03 | 0.53 | 0.96 | 0.04 | 0.44 | 0.3 |
| Age:GT | 0.48 | 0.17 | 0.79 | 1 | 0 | 0.01 | 0.3 |
| Age:WHR | 0.31 | -0.01 | 0.6 | 0.97 | 0.03 | 0.3 | 0.3 |
| Age:Haemoglobin | 0.04 | -0.28 | 0.35 | 0.59 | 0.41 | 1.81 | 0.3 |
| Age:CK | -0.35 | -0.64 | -0.04 | 0.01 | 0.99 | 0.13 | 0.3 |
| Age:LD | 0.21 | -0.07 | 0.5 | 0.93 | 0.07 | 0.72 | 0.3 |
| Age:MCHC | -0.26 | -0.54 | 0.02 | 0.04 | 0.96 | 0.41 | 0.3 |
| Age:Creatinine | -0.09 | -0.38 | 0.21 | 0.27 | 0.73 | 1.67 | 0.3 |
| Age:Magnesium | -0.14 | -0.42 | 0.13 | 0.16 | 0.84 | 1.33 | 0.3 |
| Age:Sodium | 0.02 | -0.26 | 0.28 | 0.55 | 0.45 | 2.12 | 0.3 |
| Age:Chloride | -0.07 | -0.34 | 0.2 | 0.31 | 0.69 | 1.95 | 0.3 |
| Age:MCV | 0.24 | -0.05 | 0.55 | 0.94 | 0.06 | 0.62 | 0.3 |
| Age:MCH | 0.11 | -0.18 | 0.4 | 0.76 | 0.24 | 1.46 | 0.3 |
| Age:Total_cholesterol | 0.2 | -0.08 | 0.48 | 0.91 | 0.09 | 0.88 | 0.3 |
| Age:LDL_cholesterol | 0.19 | -0.09 | 0.46 | 0.91 | 0.09 | 0.88 | 0.3 |
| Age:Systolic | 0.55 | 0.22 | 0.88 | 1 | 0 | 0.01 | 0.3 |
| Age:Diastolic | 0.22 | -0.06 | 0.55 | 0.93 | 0.07 | 0.68 | 0.3 |
| Age:Potassium | -0.04 | -0.31 | 0.23 | 0.4 | 0.6 | 1.98 | 0.3 |
| Age:Smoking | 0.35 | 0.04 | 0.67 | 0.98 | 0.02 | 0.18 | 0.3 |

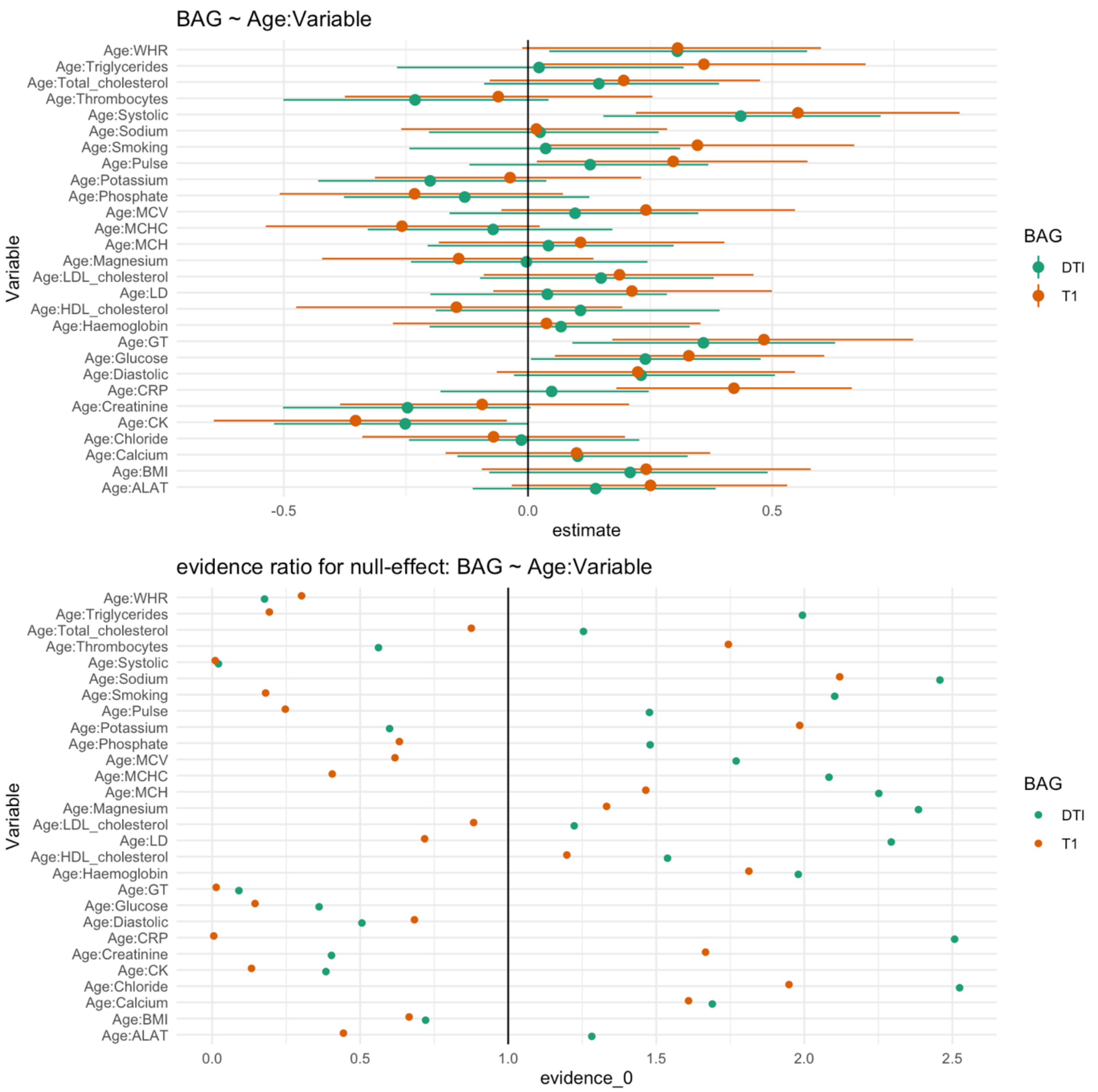


**SI Figure 17**. Estimates with 95% credible interval are shown in the upper figure and evidence ratios are shown in the lower figure. Values above 1 indicate evidence in favour of the effect being null and values below 1 indicate evidence in favour of an effect.

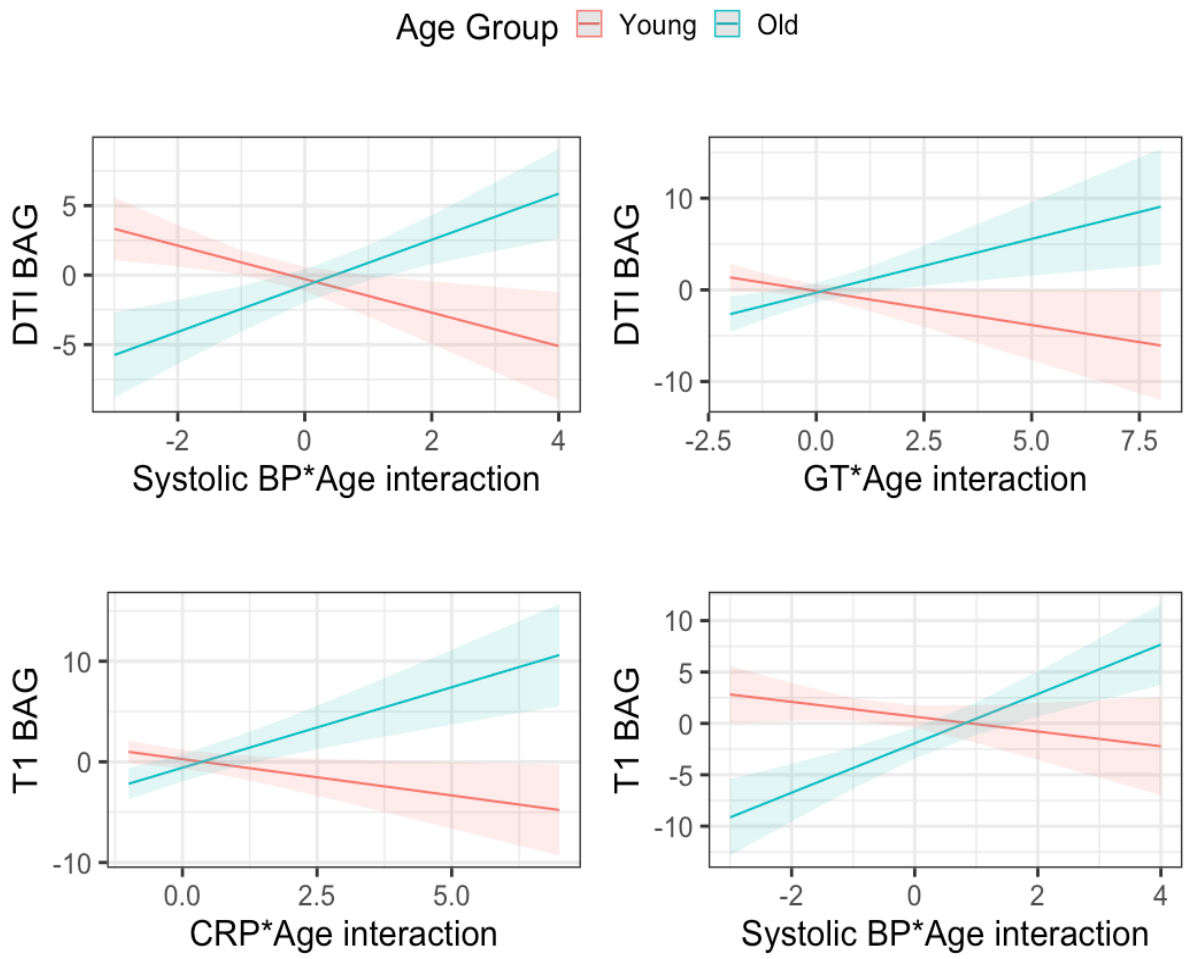


**SI Figure 18.** Interaction effects of CMH indicator variables and age on DTI and T1 BAGs, where increases in BAG are seen with increases in CMH indicator and age.
